# Mass spectrometry and machine learning for classification and molecular phenotyping of renal cell carcinoma and benign tumors

**DOI:** 10.64898/2026.01.13.26343935

**Authors:** Andreas A. Thomsen, Maj Rabjerg, Niels Marcussen, Lars Lund, Ole N. Jensen

## Abstract

Pathology classification of cancer tumor subtypes and benign tumors can be difficult due to cellular heterogeneity and similar morphological features. Renal Cell Cancer (RCC) poses a major challenge for uropathologists as advanced RCC is often incidentally diagnosed with complex histological characteristics. We hypothesised that molecular profiling of RCC and benign tumor specimen by mass spectrometry with machine learning methods enable accurate tumor classification and identification of candidate RCC biomarkers.

Mass spectrometry imaging and quantitative proteomics provided peptide and protein profiles of RCC neoplasms and benign tumors presented in tissue microarray (TMA) format of 541 tissue cores from 128 patients. Machine learning models classified RCC subtypes: Clear cell RCC (ccRCC), Chromophobe RCC (chRCC), papillary RCC (pRCC), and oncocytoma (OC). Known and novel candidate protein biomarker panels for discrimination of RCC subtypes were identified.

RCC peptide profiles and machine learning classified ccRCC, chRCC, pRCC, OC and healthy tissue with 95.6% - 100% accuracy at the patient level. Quantitative proteomics identified over 5000 proteins and machine learning classified ccRCC, chRCC, pRCC, OC and healthy tissue with 98.6% accuracy. We report new candidate protein biomarkers for ccRCC and pRCC and distinguish histologically similar subtypes OC (benign) and chRCC (malignant), as demonstrated by mass spectrometry-based immunohistochemistry. Candidate prognostic biomarkers for ccRCC contribute to risk of relapse (CDK6, CDK18) and risk of metastasis (DDB2, MK03).

Digital histo-molecular differentiation of cancer tumor subtypes and benign tumors by mass spectrometry and macbine learning enables retrospective and prospective studies. Identified candidate protein biomarkers complement existing protocols in tumor pathology.

## Introduction

Cancer remains one of the leading causes of mortality worldwide, posing immense challenges to public health and clinical treatment. Despite major advances in detection and therapy, the complex biological mechanisms driving cancer progression are still not fully understood. Renal cell carcinoma (RCC) is the 14^th^ most common cancer worldwide[1] and covers a range of cancer subtypes that are classified by their molecular traits[2]. Clear cell RCC (ccRCC) remains the dominant form of RCC accounting for 70-90% of cases. Papillary RCC (pRCC) constitutes 10-15% and chromophobe (chRCC) constitutes 3-5% of cases [3]. RCC tumor growth is often asymptomatic. In more than 60% of the cases renal masses are detected during screening for other ailments using radiological investigations such as ultrasound, computed tomography or magnetic resonance imaging (MRI) [4]. One particular challenge is the discrimination between malignant chRCC and benign oncocytoma (OC) as they both originate from the intercalated cells of the collecting ducts of the kidney[5–7], and they both contain a large number of mitochondria and often also oncocytic cells. Notably, there is no standardized immunohistochemical marker that distinguishes chRCC and OC. Furthermore, about 20% of RCC cases are diagnosed with advanced tumors and metastatic disease[8]. This complicates pathology classification because necrotic and sarcomatoid tissue have developed in advanced tumors. Thus, early detection and accurate classification of small RCC tumors is warranted [9–11]. There is a clear need for accurate biomarkers to distinguish malignant and benign RCC tumors and to detect and classify RCC tumors at an early stage.

Quantitative mass spectrometry allows for accurate comparative analysis of biomolecules in normal and perturbed cells, tissues and biofluids to identify diagnostic and prognostic biomarkers[12]. High sensitivity, mass accuracy and sample analysis rate of modern mass spectrometers permit large-scale clinical studies of biofluids and tissue samples [13]. Quantitative MS (proteomics, metabolomics) and mass spectrometry imaging have been used in clinical renal cell carcinoma-research for screening and classification purposes and for candidate biomarker discovery [14–16].

We hypothesize that mass spectrometry peptide profiling of larger numbers of human RCC specimen will generate high-information content peptide and protein datasets appropriate for machine learning models for accurate digital classification of RCC tumor subtypes. We report candidate diagnostic and prognostic protein biomarkers that discriminate RCC subtypes and we present predictive models for classification and discrimination of cancer subtypes ccRCC, pRCC, chRCC, OC and healthy control tissue.

## Methods

Renal tumor specimens were obtained from patients undergoing nephrectomy at Odense University Hospital (2006–2018) in a project approved by the Ethics Committee of the Region of Southern Denmark (Application no. 4462, Case no. S2014-0159, Protocol no. 72656). Pathologists identified representative regions containing ≥80% tumor cells, and matched adjacent normal tissue was included when available. In total, 541 needle biopsy cores (2–3 mm) from 128 patients were arranged into 26 tissue microarray (TMA) blocks, representing major renal tumor subtypes, healthy kidney, and control tissues (clinical information in Supplementary Table 1).

Formalin-fixed paraffin-embedded (FFPE) TMA sections (3 µm) were cryosectioned in duplicates for proteomics and MALDI-MSI. Sections were deparaffinized, rehydrated, subjected to antigen retrieval, and digested in situ with trypsin[17–19]. For LC-MS, peptides were extracted from individual cores and loaded onto Evotips for analysis. For MALDI-MSI peptide profiling, slides were coated with α-cyano-4-hydroxycinnamic acid. Details of MALDI matrix and spray settings are listed in Supplementary Table 2.

Targeted protein detection was performed by MALDI immunohistochemistry (MALDI-IHC) using mass-tag–conjugated antibodies (Ambergen) in selected chromophobe RCC and oncocytoma samples. Tissue processing, blocking conditions, antibody incubation, and matrix deposition followed the AmberGen MALDI-IHC workflow.

Proteomic analyses were conducted on an Evosep ONE coupled to a Bruker timsTOF Pro2 operating in diaPASEF mode. Protein identification and quantification were performed in DIA-NN using library-free searching with MaxLFQ quantification (parameters in Supplementary Table 3). MALDI-MSI was performed on a Bruker timsTOF fleX/MALDI-2 instrument at 100 µm spatial resolution, enabling acquisition of all TMA cores in <48 hours. Imaging datasets were processed in SCiLS Lab and the Cardinal R package for normalization, peak detection, alignment, and deisotoping, yielding 866 peptide features.

Statistical and computational analyses, including clustering, differential feature testing, pathway enrichment, machine learning classification, and survival analysis in ccRCC, were performed in R. A more detailed method sections can be found in Supplementary Methods.

## Results

### RCC Classification by MALDI-MSI Peptide Profiling

We hypothesized that MSI based tryptic peptide mass profiles alone, without cognate protein identification, sufficed for RCC tumor subtype separation and classification by unsupervised analysis. The MSI dataset was acquired from 26 different slides with TMAs covering 541 tissue cores. To assess data quality and integrity and to visualize the the overall trends of the dataset we performed unsupervised data analysis by hierarchical k-means clustering (Figure 2A) and dimensionality reduction by UMAP with comBAT batch correction [20] (manuscript submitted) (Figure 2B).

**Figure 1:**
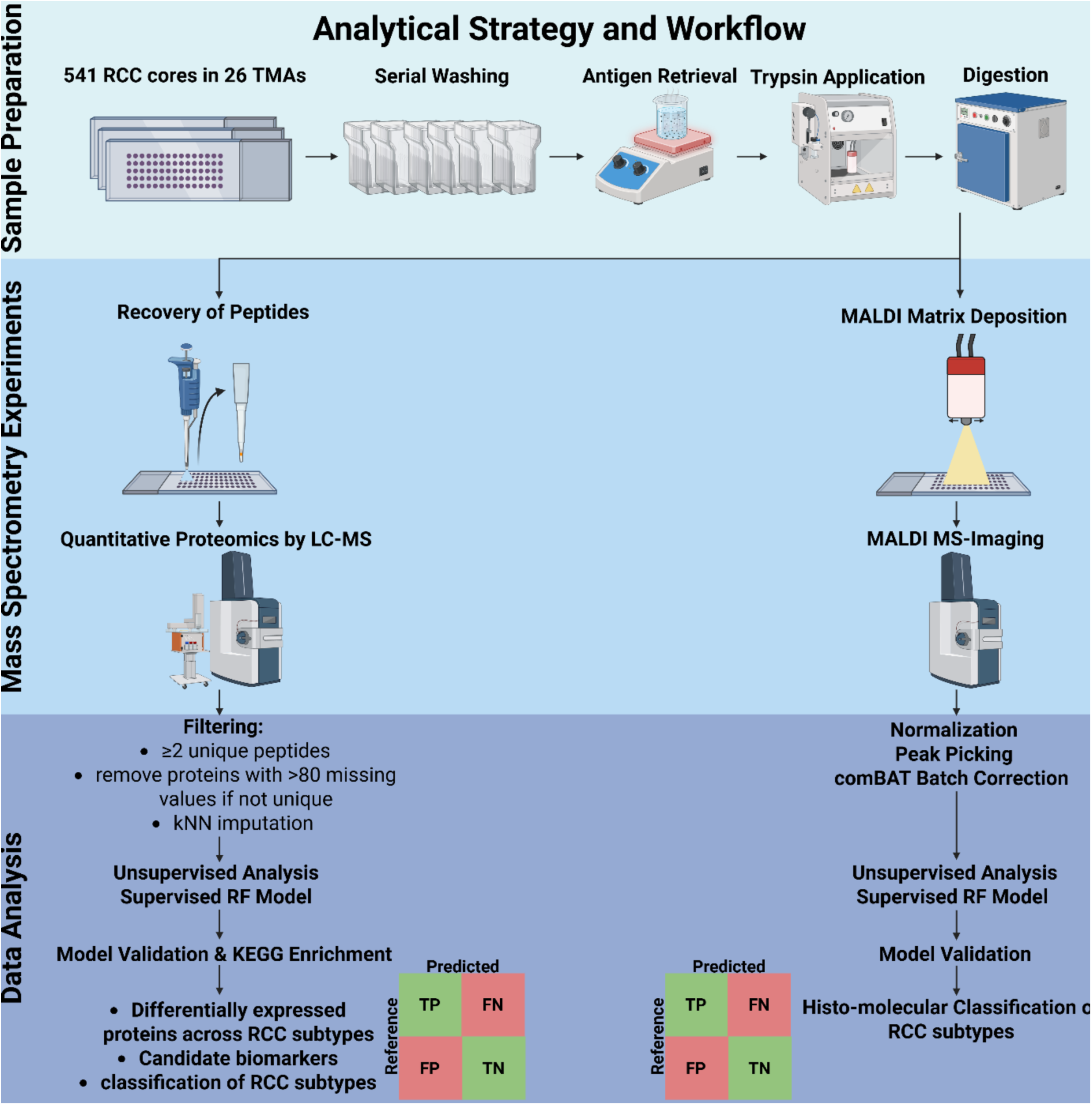
Sample preparation workflow and data analysis of combined MALDI-MSI and microproteomics datasets. TMA-FFPE blocks were sectioned in two sets: one for MSI analysis and a second for microproteomics. Both sets of samples were deparaffinized in a wash series. Subsequently samples were subdued to antigen retrieval and *in situ* trypsination. For the MSI set, matrix was applied, and samples were analyzed on a timsTOF fleX MALDI-2 system. The microproteomics samples were loaded on Evotips and run on timsTOF Pro 2 coupled to the Evosep One platform. Preprocessing of the individual MSI files were done in SCiLS Lab 2023a and imported into R v. 4.3.3 for subsequent data analysis. LC-MS files were quantified by DIA-NN v. 1.8.1. and loaded into R for subsequent data analysis. Created with Biorender.

**Figure 2:**
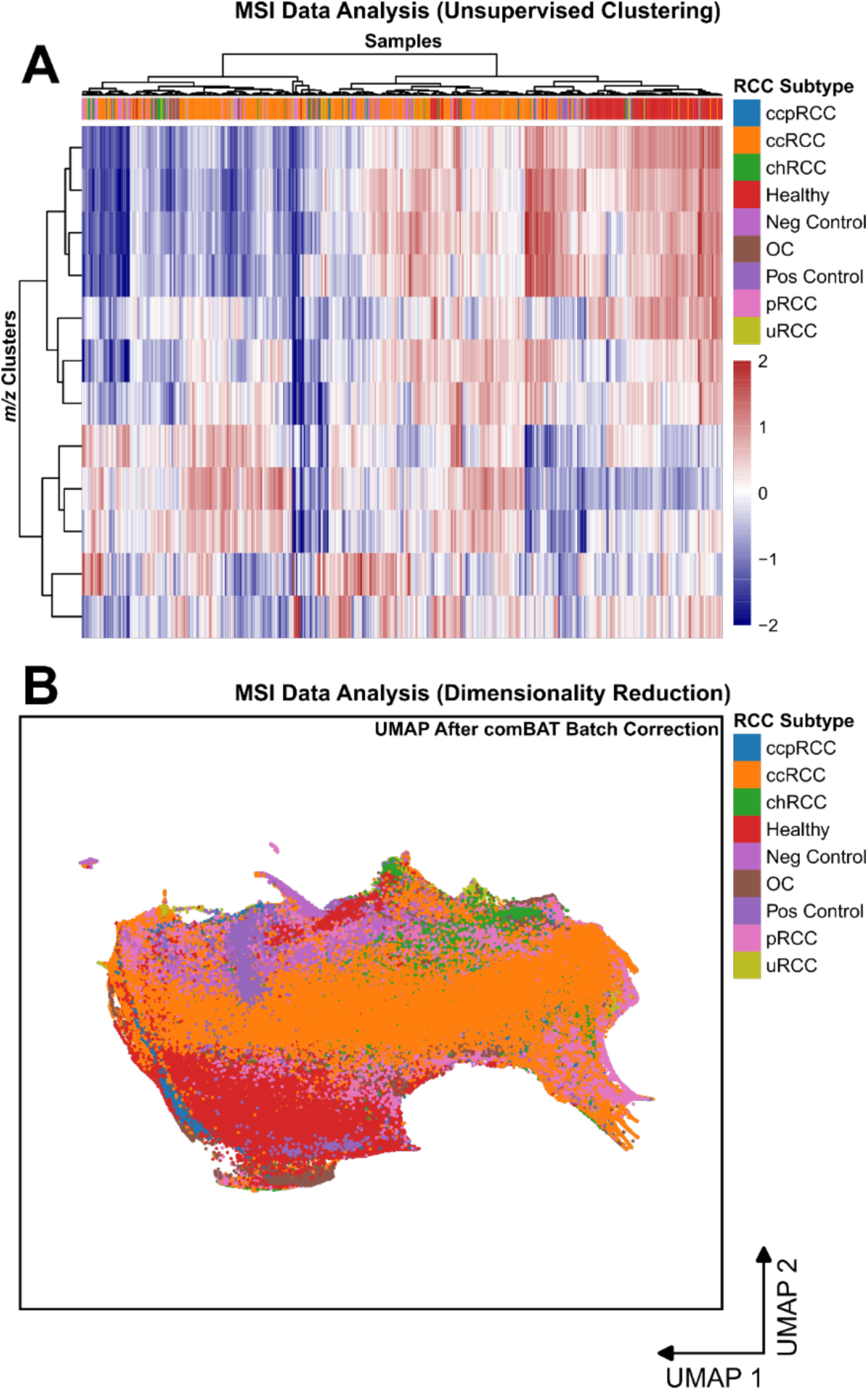
Unsupervised analysis separates RCC tumor subtypes based on peptide profiles. (A) Heatmap with 12 k-means clusters (estimated by gap statistics, within sum of squares and silhouette width) of all peptide *m/z* values on the x-axis. y-axis depicts TMA cores with every pixel belonging to one core grouped together and clustered with colors matched in RCC subtype legend. all MALDI mass spectra obtained from a single tissue core, were merged into one average sum core mass spectrum. Values are z-scores with red = higher abundance than global mean and blue = lower abundance than global mean. (B) UMAP of all pixels after comBAT batch correction. Colors indicate RCC subtype, x-axis represents first dimension of UMAP, y-axis depicts second dimension of UMAP. ccpRCC = clear cell papillary renal cell carcinoma, ccRCC = clear cell renal cell carcinoma, chRCC = chromophobe renal cell carcinoma, OC = oncocytoma, pRCC = papillary renal cell carcinoma and uRCC = unclassified renal cell carcinoma.

Unsupervised data analysis of MS imaging datasets separated the RCC subtypes and revealed similarity and homogeneity of healthy, chRCC and OC tissue cores. ccRCC and pRCC datasets were heterogenous with intra-group variation (Figure 2A). Healthy samples separate from cancerous samples. chRCC and OC cluster closely together indicating spectral overlap of the peptide profiles of these subtypes, reflecting their histo-molecular similarity. pRCC and ccRCC separate into multiple clusters with a large variance for pRCC samples. The ccRCC population exhibits a switch from low to high intensity, suggesting that additional ccRCC subgroups exist. The UMAP obtained from all 541 tissue cores after comBAT batch correction (Figure 2B) is in accordance with the hierarchical clustering analysis.

Next, we hypothesized that a machine learning model could identify distinct discriminative peptide peaks in the MSI peptide profiling data for accurate classification of RCC subtypes. We selected the random forest (RF) model because of its superior performance and ability to report features of importance as compared to other tested methods (Supplementary Figure S1). The output of the RF machine learning model for RCC classification is shown by a confusion matrix, comparing predicted diagnoses to actual diagnosis (ground truth) (Figure 3A). The RF model exhibited excellent performance for all diagnoses, and distinguished all chRCC cores from all OC cores (Figure 3A).

**Figure 3:**
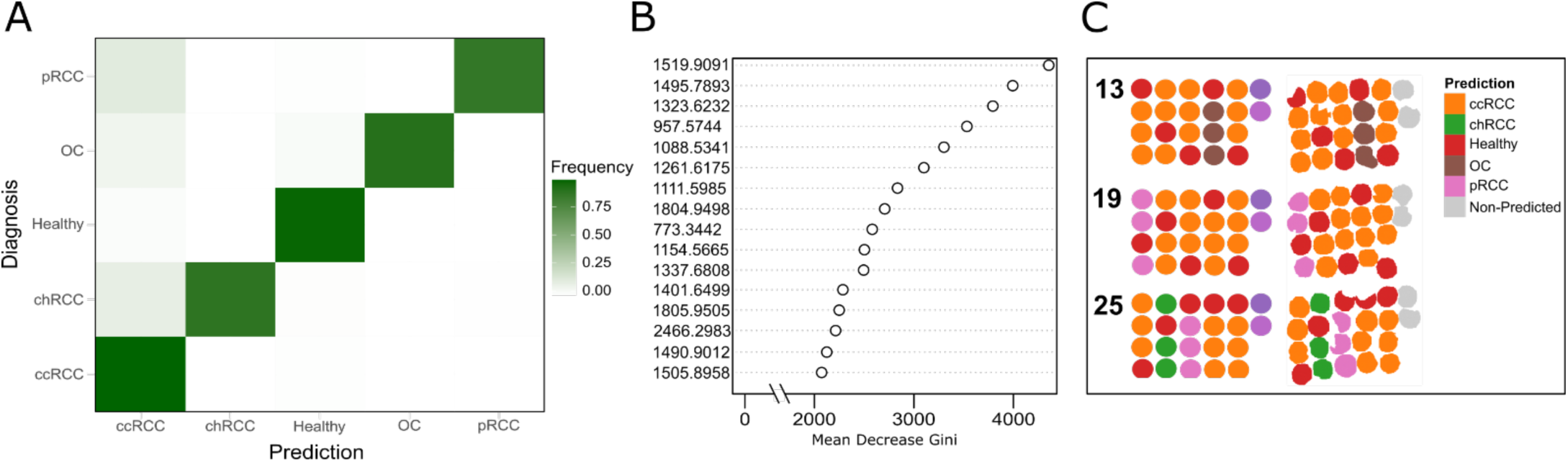
Peptide profiling by MALDI-MSI generates a highly accurate pixel-based classifier. (A) Confusion matrix based on random forest for the four RCC subtypes ccRCC, chRCC, OC, pRCC and healthy adjacent tissue. x-axis represents predicted class; y-axis represents true class. Color gradient shows relative frequency for each class. (B) variance importance plot of the top 16 ranked peptide *m/z* values according to the decrease in Gini index for the trained random forest model. x-axis represents the mean decrease in Gini index, y-axis shows *m/z* values. Circles depict the *m/z* values’ MeanDecreaseGini value. (C) Classification output on the core-level from selected TMAs. Reference cores are found on the left and predicted cores on the right. Colors indicate RCC subtypes, while grey = non-predicted are the positive and negative controls not included in the model. ccpRCC = clear cell papillary renal cell carcinoma, ccRCC = clear cell renal cell carcinoma, chRCC = chromophobe renal cell carcinoma, OC = oncocytoma, pRCC = papillary renal cell carcinoma and uRCC = unclassified renal cell carcinoma.

To test the performance of the RF model for diagnosing renal cancer at the individual patient level, we extracted the 16 most discriminative peptide mass values in the MSI data for classification of RCC subtypes across 541 cores (Figure 3B). A single tissue core was considered correctly classified if >60% of the pixels were assigned to a unique diagnosis and subsequently colored by the corresponding RCC subtype. This patient-based prediction accuracy of our RF model was 100% (Figure 3C).

### Proteome profiling of RCC tumors

We hypothesized that deep proteome analysis of 541 tumor/tissue cores by LC-MS could reveal candidate protein profiles and protein species of clinical relevance for RCC classification, diagnostics and prognostics. We identified more than 9000 proteins by LC-MS/MS based proteome analysis of the 541 RCC and tissue cores. Stringent filtering, reproducibility and quality assessment (see Suppl. Figure S2) generated a list of 5731 quantified proteins from the RCC tumors and controls.

We hypothesized that unsupervised clustering of the proteomics dataset obtained from the TMA cores could stratify the RCC subtypes and reveal biological and pathological processes within each RCC subtype. All RCC subtypes form distinct clusters, with ccRCC exhibiting two protein subgroups, defined by clusters 8 and 12 (Figure 4A). The two tissue control groups cluster separately, indicating distinct protein expression patterns relative to RCC tumor subtypes. Six protein clusters (clusters 4, 5, 6, 7, 8, and 12) exhibited discriminating patterns for RCC subtypes and were further analysed (Figure 4B). As expected, chRCC and OC exhibited similar protein expression profiles, with chRCC showing higher protein abundance in cluster 4 and OC displaying higher protein abundance in cluster 5.

**Figure 4:**
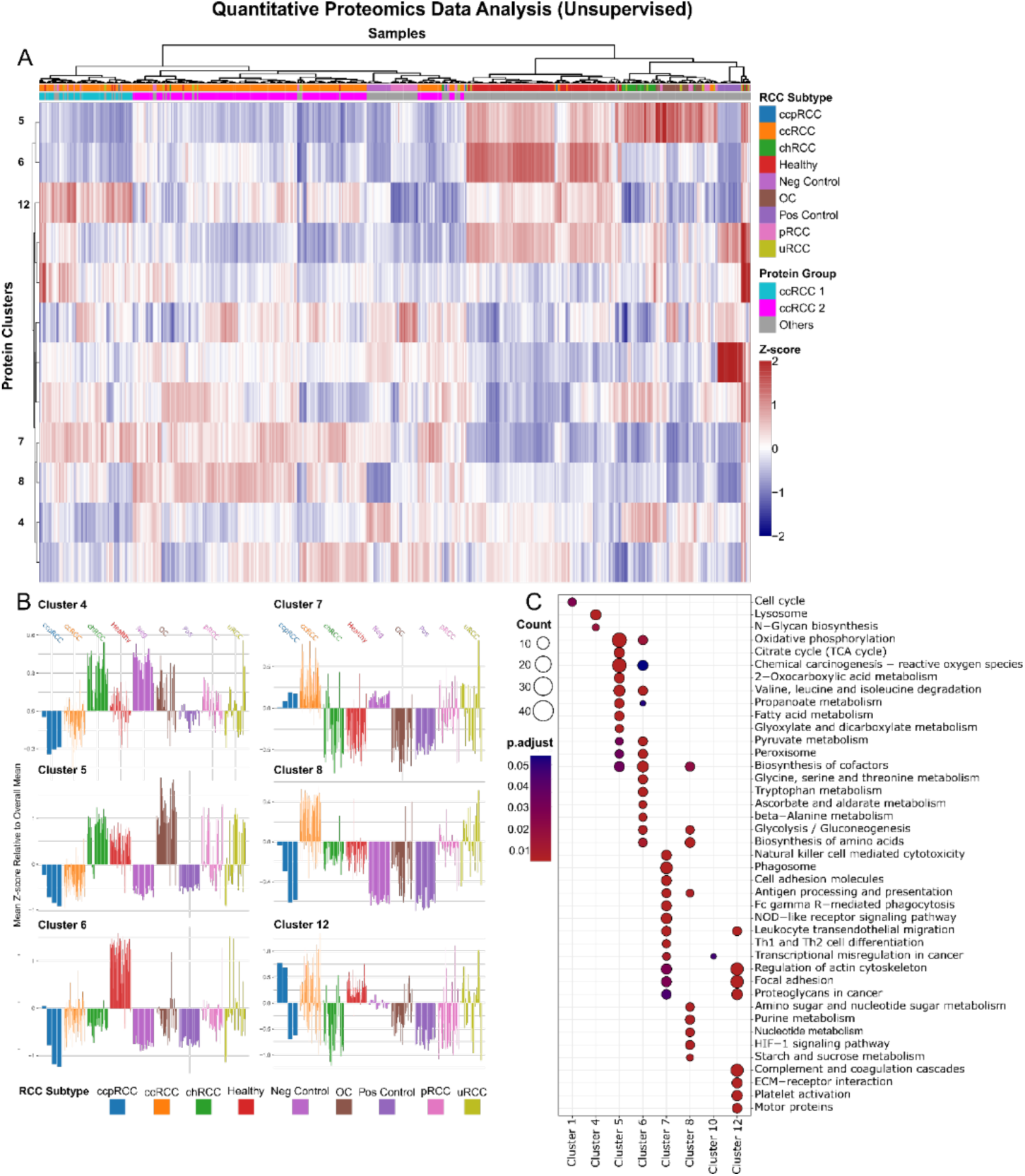
Unsupervised analysis and enrichment provide distinct phenotypes of RCCs and OC. (A) Heatmap with 12 k-means clusters of all proteins on the x-axis. y-axis depicts samples clustered with colors matched in RCC subtype legend and with ccRCC protein groups. Values are z-scores with red = higher abundance than global mean and blue = lower abundance than global mean. Clusters of interest are labelled on the x-axis. (B) Barplots of isolated protein clusters 4, 5, 6, 7, 8, and 12 from heatmap in panel A. Each bar represents one sample and is colored according to RCC subtype legend. X-axis is bars, y-axis is mean z-score relative to global mean. (C) Dotplot of cluster comparison of enriched KEGG terms found in all the protein clusters from panel A at p and q-value cutoff of ≥0.05 adjusted with Benjamini-Hochberg. Count of proteins is depicted as circle size in legend, adjusted p-value is indicated by color in legend. x-axis represents clusters, y-axis represents KEGG terms. ccpRCC = clear cell papillary renal cell carcinoma, ccRCC = clear cell renal cell carcinoma, chRCC = chromophobe renal cell carcinoma, OC = oncocytoma, pRCC = papillary renal cell carcinoma and uRCC = unclassified renal cell carcinoma.

Biochemical pathway analysis of the proteomes by KEGG enrichment analysis revealed molecular and cellular functions of the identified protein clusters of RCC (Figure 4C). For example, protein cluster 4 was enriched for lysosomal proteins, while cluster 5 contained mitochondria-related proteins, e.g., citrate cycle, fatty acid metabolism, glyoxylate, and dicarboxylate metabolism (Figure 4C). Angiogenesis-related terms were prominent in cluster 12.

KEGG analysis indicated that chRCC and OC contain high levels of mitochondrial proteins relative to other RCC subtypes and normal tissue. ChRCC displayed elevated levels of lysosomal proteins, e.g. lysosome-associated membrane proteins 1 and 2 (LAMP1/2). All ccRCC samples exhibited high levels of immune-related KEGG terms (clusters 7 and 8), indicating a role of the tumor microenvironment (TME) in the clear cell RCC phenotype. ccRCC comprises two distinct protein groups defined by clusters 8 and 12, that stratifies samples into ccRCC2 and ccRCC1 groups. ccRCC1 may promote metastasis through EMT due to higher levels of laminins involved in basement membrane degradation[21], integrins (Integrin αV and Integrin β1) involved in cell adhesion[22] and myosins (Myosin-9 and −10) linked to cell motility[23]. The ccRCC2 protein group seems metabolically adapted towards the Warburg effect with more abundant glycolytic proteins [24–26]. while also being hypoxia-responsive [27]. The most defining KEGG terms associated with each subtype can be found in Supplementary Table 4.

In summary, quantitative proteome analysis of RCC subtypes followed by KEGG enrichment identified biological processes and molecular phenotypes specific for distinct RCC subtypes. The proteome of the ccRCC subtype was strongly enriched in immune-related proteins involved in innate and adaptive immunity, respectively. This suggests that ccRCC activates the immune response that in turn affects the tumor microenvironment (TME). These immune system activation and infiltration features were not observed in any of the other RCC subtypes. Pairwise comparison across the investigates RCC subtypes (Supplementary Figure S3), is consistent with the results shown in Figure 4C.

### RCC proteome data correlates with current clinical protocols

Next, we asked whether the quantitative proteomics datasets included currently used biomarkers for RCC diagnostics, i.e. immunohistochemistry panels, for ccRCC, pRCC, chRCC, and OC (Table 1). Indeed, all four proteins currently used in IHC assays for RCC subtype classification were identified and quantitified (upregulated) in our large-scale proteomics dataset (Table 1). We therefore hypothesized that the proteomics datasets obtained from the RCC subtypes also include novel candidate protein biomarkers suitable for precise diagnosis and prognosis of RCC tumors. To identify such features we trained a random forest classifier on the complete RCC proteomics data to discriminate RCC subtypes (Figure 5). The confusion matrix displays a near perfect RCC subtype prediction on the test set with a mean balanced accuracy of 98.6% (Figure 5A). Next, we visualized the similarity level across the predicted test set samples to reveal how the model perceives the data. To do so we performed multi-dimensional scaling for the predicted test set (Figure 5B). Whereas OC and chRCC sample sets cluster closely together, the RF model perfectly distinguishes between these two subtypes (Figure 5B).

**Figure 5:**
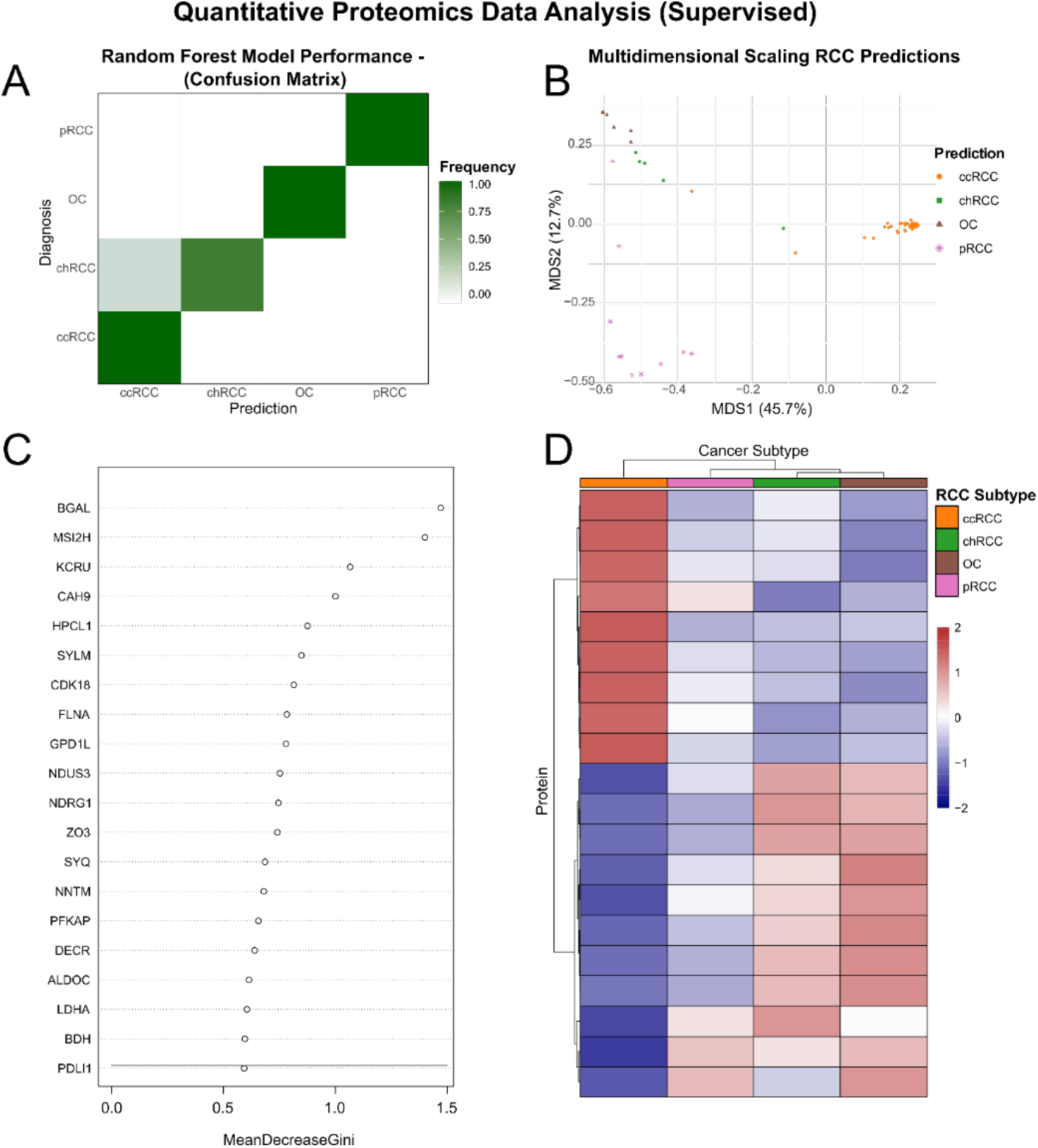
The random forest algorithm proposes novel diagnostic protein biomarkers. (A) Confusion matrix based on random forest for the four RCC subtypes ccRCC, chRCC, OC and pRCC. x-axis represents predicted class; y-axis represents true class. Color gradient shows relative frequency for each class. (B) Multi-dimensional scaling (MDS) of predicted microproteomics sample. Each symbol is a predicted sample within the multi-dimensional space. X-axis represents the first dimension (explains 45.7% of predicted data variance) of the MDS, y-axis represents second dimension (explains 12.7% of the predicted data variance) of the MDS. (C) variance importance plot of the top 20 ranked proteins according to the decrease in Gini index for the trained random forest model. x-axis represents the mean decrease in Gini index, y-axis shows Uniprot protein entries. Circles depict the Uniprot entries’ MeanDecreaseGini value. (D) Heatmap of the top 20 ranked proteins found in 4C. The proteins are clustered on the y-axis while the x-axis shows the clustering of mean of RCC subtypes included in the random forest model. Values are z-scores with red = higher abundance than global mean and blue = lower abundance than global mean. ccRCC = clear cell renal cell carcinoma, chRCC = chromophobe renal cell carcinoma, OC = oncocytoma and pRCC = papillary renal cell carcinoma.

**Table 1:** Microproteomics data correlates with immunohistochemistry panel. Standard panel of renal cell carcinoma immunohistochemistry proteins in Denmark. Rows are proteins, four first columns are subtypes, entries indicate the positivity level of the stain with +++ = highly positive and - = absent. Cytokeratin 7 (K2C7), vimentin (VIME), carbonic anhydrase 9 (CAIX) and alpha-methylacyl-CoA racemase (AMACR) are used as the routine standard panel in pathology departments in Denmark. Fifth column depicts violinplots with relative protein abundance across RCC subtypes for the four standard panel proteins. y-axis represents the relative abundance log2-scale. p-values are calculated by Mann-Whitney Test with significance levels: NS. = >0.5, * = ≤0.05, ** = ≤0.01, *** = ≤0.001. ccRCC = clear cell renal cell carcinoma, chRCC = chromophobe renal cell carcinoma, OC = oncocytoma and pRCC = papillary renal cell carcinoma.

|  | ccRCC | chRCC | OC | pRCC | Quantitative Proteomics |
| --- | --- | --- | --- | --- | --- |
| K2C7 | - | +++ | Rare | +++ |  |
| VIME | +++ | - | - | +++ |  |
| CAIX | +++ | - | - | - |  |
| AMACR | - | - | - | +++ |  |

Next, we identified novel potential protein biomarkers for RCC subtypes by extracting features from the RF model (Figure 5C). A striking observation is the high ranking of Carbonic Anhydrase 9, an established biomarker component of the standard immunohistochemistry panel employed by pathologists to differentiate RCC subtypes (discussed below). Other proteins with distinct roles in cancer included NDRG1, Programmed Death Ligand 1, Cyclin Dependent Kinase 18, Filamin-A and RNA-binding protein Musashi homolog 2 [28–32].

We evaluated the discriminatory power of these proteins and their expression levels across the RCC subtypes by hierarchical clustering (Figure 5D). One protein cluster is characterized by upregulation in ccRCC and downregulation in the remaining subtypes, and another protein cluster is highly expressed in chRCC and OC, and partly in pRCC.

Our RCC proteome analyses allowed us to propose a panel of candidate diagnostic protein biomarkers for each RCC subtype (Table 2). Candidate proteins for ccRCC are linked to the TME including immune infiltration and the Warburg effect. chRCC biomarker candidates are linked to vesicle transport and lysosomes whereas OC protein candidates are found in the mitochondria. pRCC markers include proteins with various functions related to cancer.

**Table 2:**
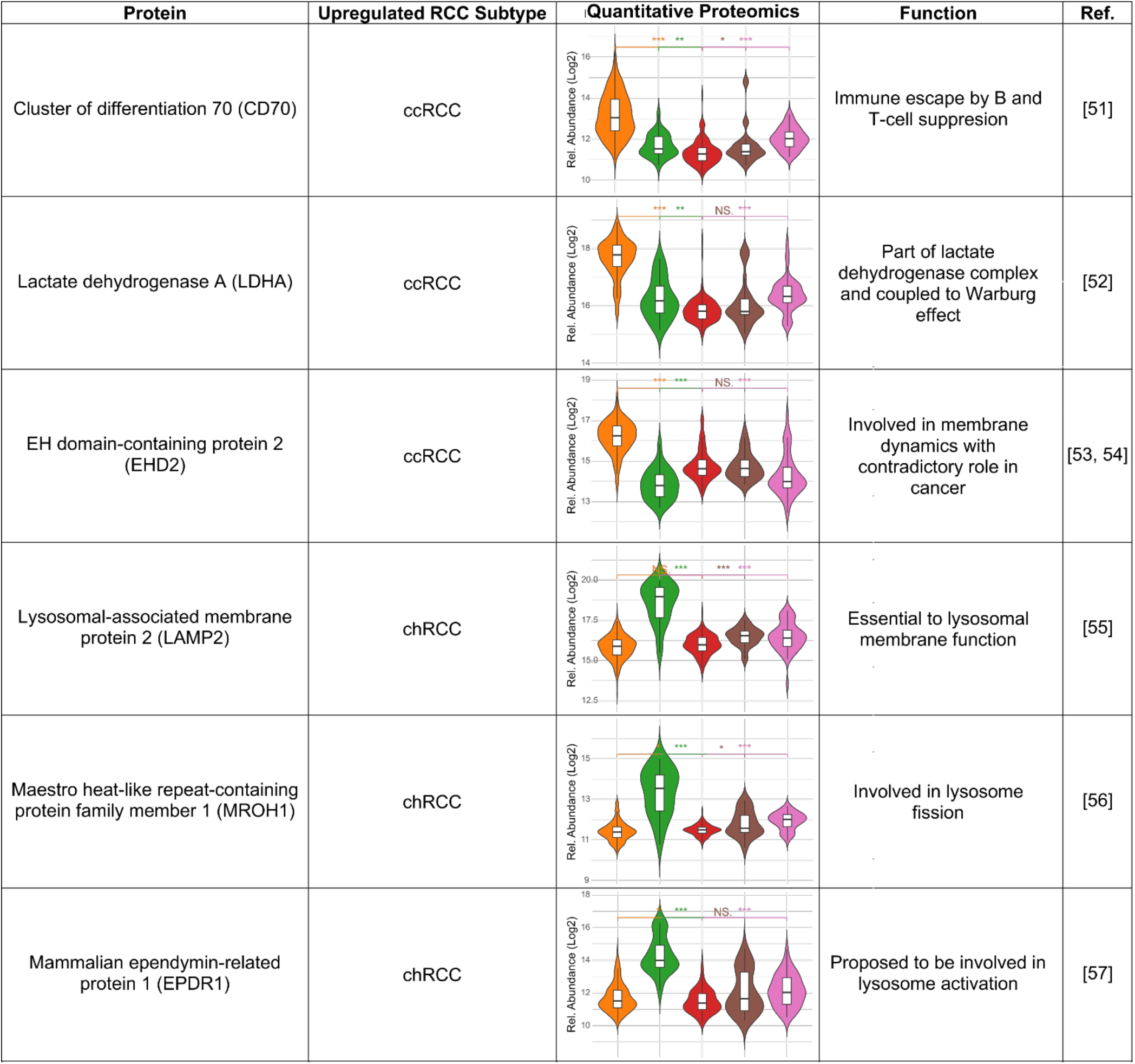
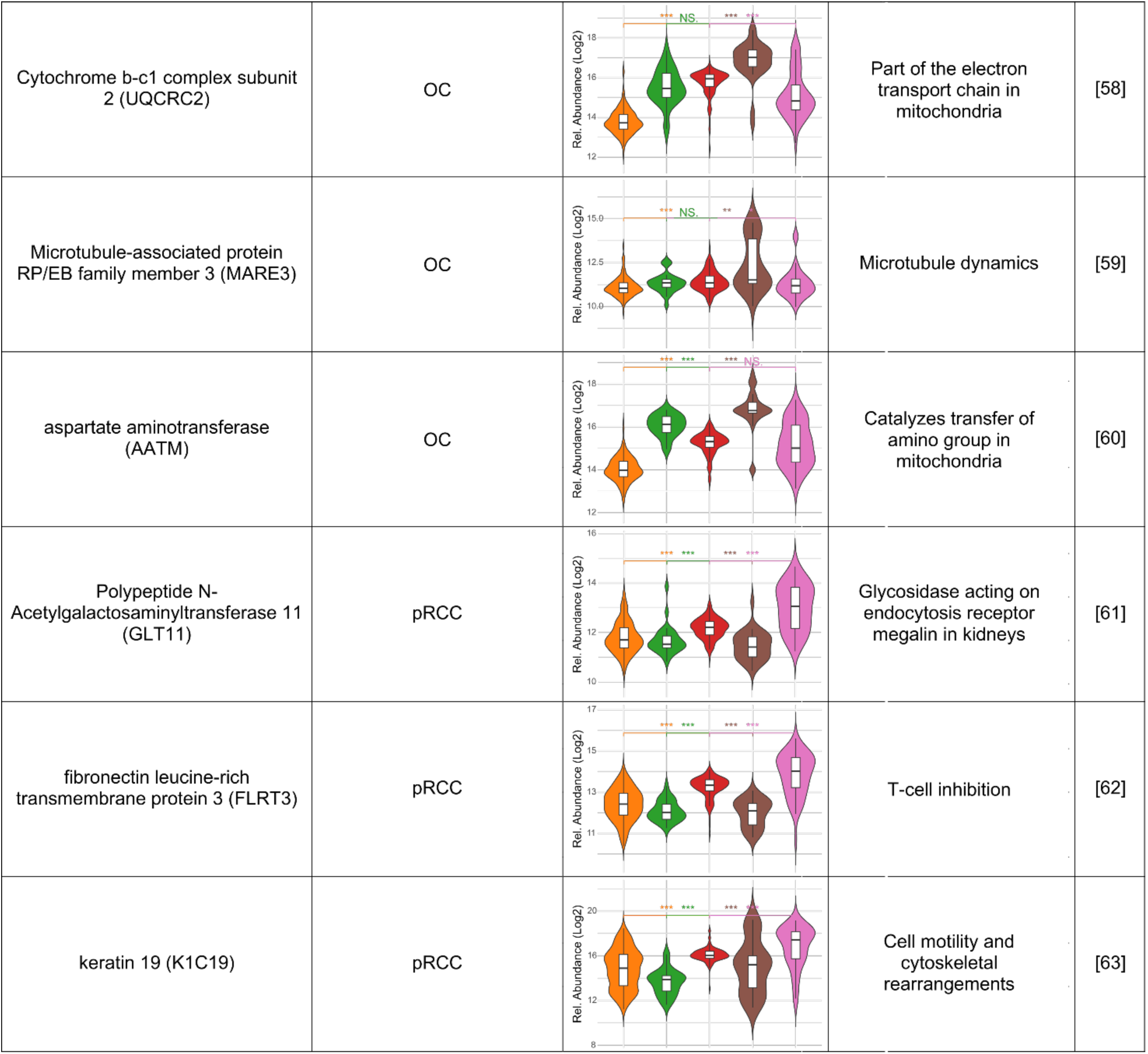
Novel proposed diagnostic protein biomarkers’ function and relative abundance. Functions of proposed novel diagnostic protein biomarkers known from literature. Rows are proteins. The first columns are protein name, which subtype it was found at higher levels and its’ function. Fifth column depicts violinplots with relative protein abundance across RCC subtypes for the proposed candidates. y-axis represents the relative abundance log2-scale. p-values are calculated by Mann-Whitney Test with significance levels: NS. = >0.5, * = ≤0.05, ** = ≤0.01, *** = ≤0.001. ccRCC = clear cell renal cell carcinoma, chRCC = chromophobe renal cell carcinoma, OC = oncocytoma and pRCC = papillary renal cell carcinoma.

In summary, the RCC-proteome trained random forest model achieved near-perfect RCC tumor classification and effectively identified candidate biomarkers for RCC subtypes.

### Relapse Analysis and Leibovich Risk Assessment Identifies Prognostic Protein Biomarkers

We used the extensive clinical metadata from ccRCC patient records (n = 83) to identify prognostic protein biomarkers for the ccRCC group and to predict relapse probability and metastasis risk. First, ccRCC patients were categorized into two groups: a high protein level group (>1.05× median) and a low protein level group (<0.95× median). Using Cox regression analysis, we assessed whether protein levels influenced relapse probability. A hazard ratio (HR) > 1 indicated that low protein expression corresponds to a worse prognosis, while HR < 1 is indicative of a better prognosis. Significant associations between relapse probability and protein levels were identified (Figure 6A). Further, Kaplan-Meier survival curves were generated for the high and low expression groups of two key proteins: cyclin-dependent kinase 6 (CDK6) and ectonucleotide pyrophosphatase/ phosphodiesterase family member 3 (ENPP3) (Figure 6B-C). Both proteins were found to be significant in the Cox regression test and exhibited consistent trends in the Kaplan-Meier curves, confirming their potential as prognostic biomarkers.

**Figure 6:**
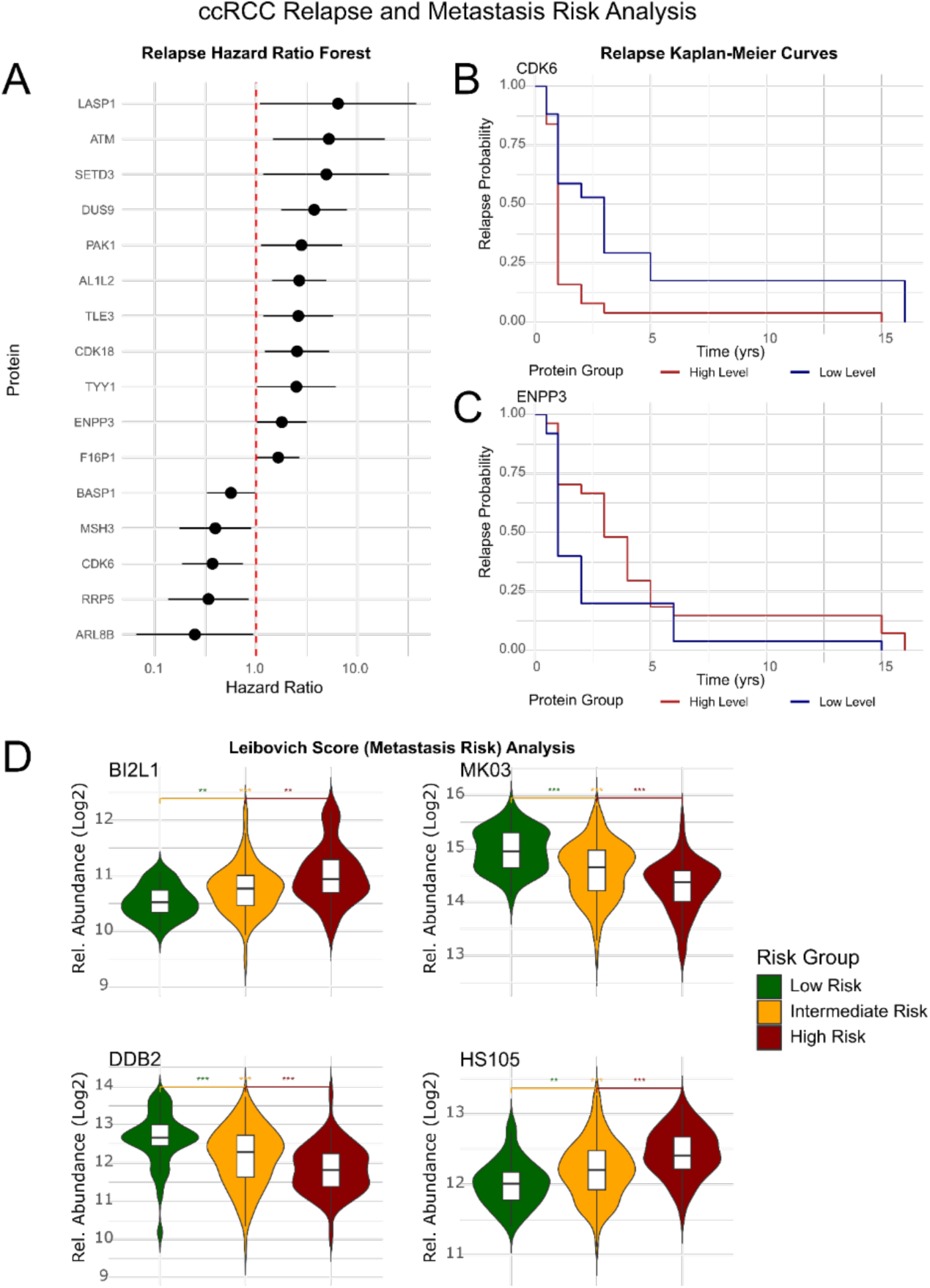
Relapse and metastasis risk analysis for ccRCC subtype proposes prognostic protein biomarkers. (A) Hazard ratio forest plot of significant cancer-associated proteins. x-axis represents hazard ratio; y-axis depicts protein names. Dashed red line is at HR = 1 which indicates a change in prognosis. Each proteins mean value is at the circle with lines indicating confidence intervals. (B) Kaplan-Meier curves for CDK6 high protein level = red vs low protein level = blue groups. x-axis represents time in years; y-axis depicts relapse probability. (C) Kaplan-Meier curves for ENPP3 high protein level = red vs low protein level = blue groups. x-axis represents time in years; y-axis depicts relapse probability. (D) violinplots of BI2L1, MK03, DDB2 and HS105 for the Leibovich risk groups: Low risk = 0-2, intermediate risk = 3-5, high risk = ≥6. y-axis represents the relative abundance log2-scale. p-values are calculated by Mann-Whitney Test with significance levels: NS. = >0.5, * = ≤0.05, ** = ≤0.01, *** = ≤0.001. ccRCC = clear cell renal cell carcinoma, chRCC = chromophobe renal cell carcinoma, OC = oncocytoma and pRCC = papillary renal cell carcinoma.

Next, we incorporated Leibovich scoring to stratify ccRCC patients into three distinct risk groups (low risk = 0–2, intermediate risk = 3–5, high risk = >6). Statistical testing revealed that relative protein levels of BAR/IMD domain-containing adapter protein 2-like 1 (BI2L1), Mitogen-activated protein kinase 3 (MK03), DNA damage-binding protein 2 (DDB2), and Heat shock protein 105 kDa (HS105) varied significantly across risk groups (Figure 6D). Specifically, B12L1 and HS105 levels increased progressively with higher risk, while MK03 and DDB2 levels decreased with increasing risk of metastasis. In summary, we identified several candidate prognostic protein biomarkers for ccRCC that provides a foundation for further studies exploring their clinical utility for predicting relapse and metastasis risk.

### Validation of Diagnostic Protein Biomarkers by MALDI Immunohistochemistry (MALDI-IHC)

We confirmed two candidate biomarkers using a novel immunohistochemistry method: Coupling of MALDI-MSI and immunohistochemistry (IHC) by using antibodies conjugated with photocleavable mass tags, known as MALDI-IHC [46]. We aimed to discriminate the clinically challenging malignant chRCC tumor from the benign OC tumor utilizing primary rabbit antibodies targeting human LAMP2 (chRCC) and UQCRC2 (OC) (Figure 7). The MALDI-IHC image readout of anti-UQCRC2 antibody (Figure 7A, left) demonstrated a strong signal intensity in OC tissue sections specific to the affected tissue area, whereas the anti-LAMP2 antibody (Figure 7A, right) exhibited high intensity in chRCC tumor tissue sections, but not in adjacent healthy parenchyma. We then performed MALDI-IHC analysis in TMA format of RCC cancer subtypes including chRCC and OC patients (TMA#8). The anti-UQCRC2 antibody produced signals in OC cores (Figure 7C) and the anti-LAMP2 antibody generated signals in chRCC cores (Figure 7D), and not vice versa. Quantitative analysis of signal intensity of the ChRCC tumors or OC masses (Figure 7E-F) displays a statistically significant difference, demonstrating that chRCC tissue exhibited higher levels of LAMP2 compared to OC tissue, while OC tissue had higher levels of UQCRC2 compared to chRCC cores. These results support the hypothesis that one or few distinct proteins may serve as reliable diagnostic biomarkers for discriminating chRCC and OC.

**Figure 7:**
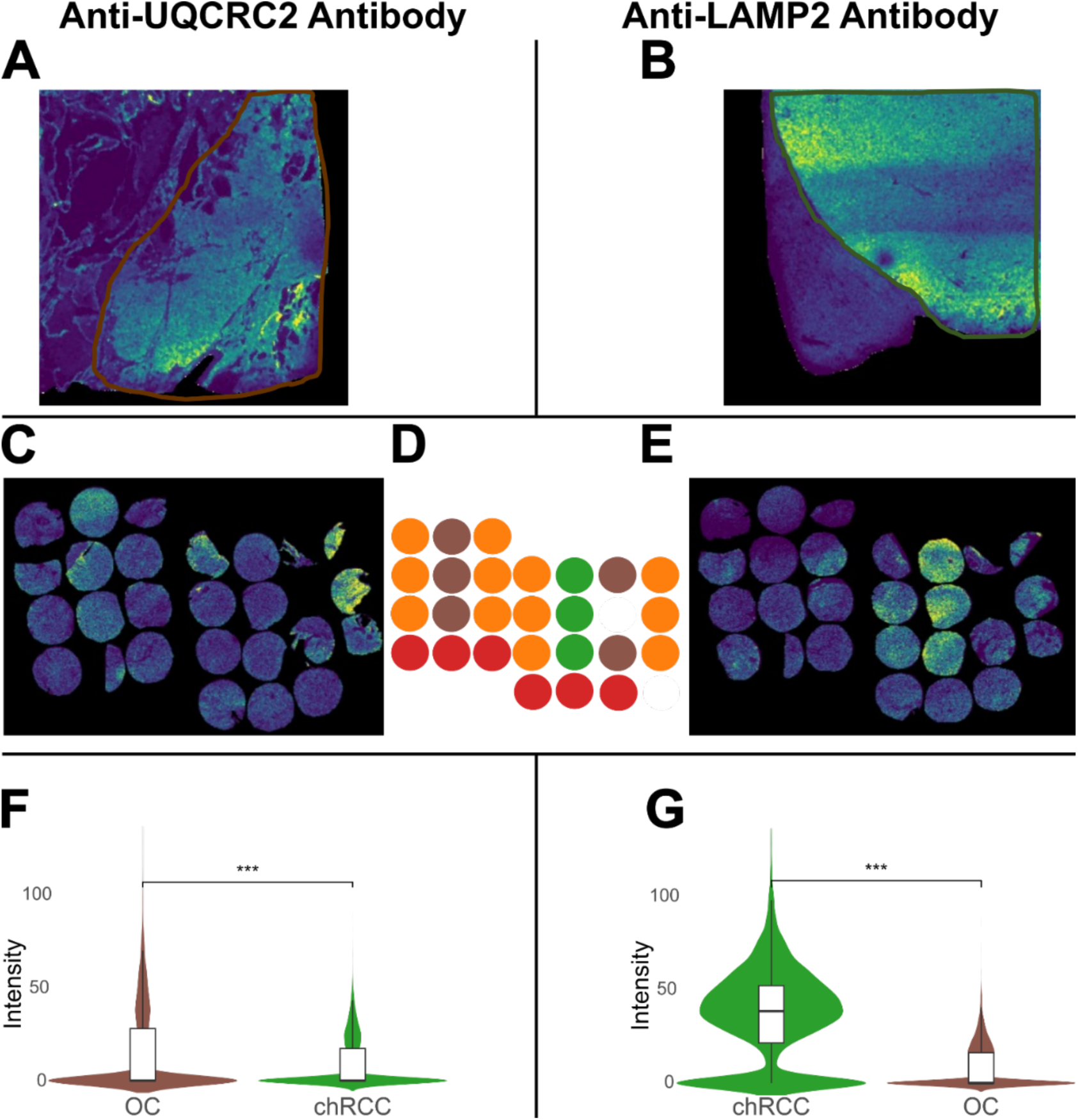
Validation by MALDI-IHC confirms novel diagnostic protein biomarkers. (A) Ion image of LAMP2 secondary antibody mass tag from large chRCC tissue section. Scale is 4 mm. (B) Ion image of UQCRC2 secondary antibody mass tag from large OC tissue section. Scale is 4 mm. (C) Reference RCC subtype for selected tissue microarray found in C and D. (C) ion image of anti-UQCRC2 secondary antibody mass tag from selected tissue microarray. (D) ion image of anti-LAMP2 secondary antibody mass tag from selected tissue microarray. (E) Intensity box and violin plot of OC and chRCC tissue cores from selected tissue microarray for all pixels. y-axis depicts normalized intensites, p-values are calculated by Wilson-Cox Test with significance levels: NS. = >0.5, * = ≤0.05, ** = ≤0.01, *** = ≤0.001. (F) Intensity box and violin plot of chRCC and OC tissue cores from selected tissue microarray for all pixels. p-values are calculated by Mann-Whitney Test with significance levels: NS. = >0.5, * = ≤0.05, ** = ≤0.01, *** = ≤0.001.

## Discussion

Our comprehensive peptide and protein profiling of RCC and OC tissue cores by two complementary mass spectrometry technologies, MALDI MS and ESI MS, generated large-scale datasets for automated digital RCC tumor classification by machine learning and for molecular characterization of perturbed protein networks in RCC subtypes.

The advantage of MALDI MS for peptide profiling is the simple and semi-automated sample preparation, high mass accuracy for peptide profiling, and its data acquisition speed. MS results from the 541 RCC cores were obtained in less than 48 hours of data acquisition. The information content of the spectral peptide mass profiles in the range m/z 600-3500 suffices to classify the RCC subtypes without the need for protein identification. Importantly, the model can distinguish between chRCC and OC with no misclassifications, which has potential clinical impact due to the current challenges in distinction of these two tumor subtypes. The model could serve as a second opinion to aid the uropathologists when classifying renal cancers and prevent avoidable surgical intervention. The few misclassified samples (pixels) might be due to an imbalanced dataset containing a majority of ccRCC cores. To avoid misclassification, more samples of rare non-ccRCC tumors should be included in the training set. Nevertheless, the model perfectly discriminates the included tumor subtypes at the patient-level highlighting the clinical utility of MSI.

MALDI-MSI combined with computational classification has previously been used to differentiate RCC subtypes. Earlier studies reported subtype discrimination using lipid or metabolite MSI profiles with PLS-DA, SVM, or Lasso models, achieving accuracies ranging from ∼77–99% depending on subtype pairs and molecular class[47–52]. We similarly separated ccRCC, chRCC, and OC using MSI peptide profiling (87% accuracy) and achieved 100% accuracy with LC-MS proteomics and an SVM model, highlighting the complementary strengths of MSI and LC-MS approaches.

In the present study, we analyzed all common malignant RCC subtypes, OC, and healthy tissue, representing, to our knowledge, the largest and most diverse MSI-based RCC classification dataset to date. A random forest model trained on 541 peptide mass profiles achieved a weighted mean balanced pixel-level accuracy of 95.6%. The use of a random forest classifier was advantageous not only for performance but also for providing feature importance ranking, facilitating identification of discriminatory peptide and protein signals while reducing the risk of overfitting.

Our new quantitative proteomics workflow based on *in situ* digestion of TMA cores and DIA-PASEF LC-MS/MS was adapted from our previous study [50]. To automate the sample preparation and to avoid cross-contamination (by peptide diffusion) of the tissue cores we choose to perform digestion by spraying the trypsin solution instead of manually positioning trypsin droplets. The identification of over 9000 proteins and more than 5000 unique proteins on average across all samples and RCC subtypes (supplementary Figure 2B) showcases the high performance of current proteomics technology. We identified and quantified all four biomarker proteins used in current RCC diagnostics, also matching the expected protein levels (Table 1). We report a list of novel candidate protein biomarkers for distinguishing ChRCC and OC (Table 1).

Clear cell RCC has been widely characterized at the transcriptomic and proteomic levels, and large-cohort studies consistently highlight its pronounced immunogenic and angiogenic phenotype relative to normal kidney, pRCC, and chRCC[14, 53, 54] In agreement, we observed increased leukocyte infiltration, antigen processing, and HIF1-driven angiogenesis in ccRCC (Supplementary Fig. 3D). ccRCC also exhibited metabolic reprogramming with increased monosaccharide utilization and reduced TCA/OXPHOS activity, consistent with a Warburg-like shift[50, 53, 55], together with enhanced glutamine metabolism supporting aberrant lipid synthesis, a hallmark of this subtype[56], (Supplementary Figure 3A) and upregulated glutathione metabolism, potentially providing protection against ferroptosis[57, 58]. Moreover, ccRCC showed elevated expression of proteins involved in cytoskeletal remodeling, focal adhesion, ECM-receptor interaction, and Rap1 signaling, indicating a higher epithelial–mesenchymal transition phenotype and increased metastatic readiness compared with other subtypes (Supplementary Fig. 3C) [59, 60]. Additionally, ccRCC samples separated into two proteomic groups: one more hypoxia-adapted and metabolically reprogrammed, and another more invasive (Fig. 4A–C), consistent with previous findings[16].

In contrast, both oncocytoma and chromophobe RCC relied more on oxidative metabolism, reflected in increased mitochondrial pathway activity (Supplementary Figure 3A). In chRCC, elevated mitochondrial genome copy number and enhanced TCA/OXPHOS flux have been linked to tumor growth[61], In OC, defective complex I activity and reduced HIF1A result in the accumulation of structurally abnormal mitochondria with high protein abundance but limited oxidative capacity [62, 63]. Direct comparison of the two subtypes revealed higher apparent mitochondrial metabolic protein levels in OC, whereas chRCC was characterized by increased endocytosis and lysosomal trafficking pathways[50, 64, 65]. AMPK signaling, a positive regulator of mitochondrial biogenesis, was uniquely elevated in chRCC (Supplementary Fig. 3B)[54]. In contrast, pRCC did not display distinct pathway enrichment patterns, likely reflecting the molecular heterogeneity of type I and II tumors.

Comparative quantitative proteomics defined molecular protein profiles of the RCC subtypes. They included presently used clinical RCC classification biomarkers and identified a panel of novel candidate diagnostic biomarkers. We validated two of these candidates (LAMP2 and UQCRC2) based on their clinical relevance in discriminating malignant chRCC tumors from benign OCs by MALDI-IHC (Figure 7). This confirms the depth and quality of our quantitative proteomics dataset and highlights the utility of mass spectrometry-based immunohistochemistry for independent verification.

Previous studies reported S100 calcium binding protein A1 (S100A1), hepatocyte nuclear factor 1 beta (HNF1β), cytokeratin 7 (K2C7), caveolin-1, lysosomal-associated membrane protein 1 (LAMP1) and integrin alpha V (ITGAV) as candidate diagnostic biomarkers distinguishing OC from chRCC[65–67]. However, these proteins were not translated into clinical use due to overlapping staining. Interestingly, LAMP1 and LAMP2 (our validated candidate) are closely related with the same functions in the lysosomal membrane, which led us to search for related proteins. The study from Drendel et al[65] mentions LAMP3/CD63 as discriminatory and we observe significantly higher relative abundances of both LAMP1 and LAMP3 in chRCC compared to OC (not shown). A more recent CPTAC study highlighted new markers, microtubule associated protein RP/EB family member 3 (MARE3), transmembrane glycoprotein NMB (GPNMB) and adhesion G Protein-coupled receptor F5 (ADGRF5)[68]. The CPTAC study and our results reveal higher levels of MARE3 protein in OC compared to chRCC, so this candidate should be further explored.

Ataxia telangiectasia mutated (ATM), a key regulator of the DNA damage response, has been linked to poor survival when downregulated [69, 70], which is consistent with our observation that low ATM protein levels were associated with increased relapse risk (Figure 6A). Similarly, reduced levels of SETD3, a histidine methyltransferase promoting apoptosis after DNA damage [71, 72], was also associated with higher relapse probability in ccRCC.

Two cyclin-dependent kinases also contributed to relapse risk. Low CDK6 levels were linked to better outcomes, supporting ongoing interest in CDK6 inhibition as a therapeutic approach in RCC[73]. In contrast, high CDK18 levels were associated with reduced relapse risk, in line with findings in triple-negative breast cancer[28].

Low LIM and SH3 domain protein 1 (LASP1) expression correlated with increased relapse risk in our cohort; however, prior studies report conflicting prognostic associations[72, 74], underscoring the need for further evaluation using quantitative proteomics.

Leibovich risk group analysis (Figure 6D) identified several proteins associated with metastasis likelihood. BAR/IMD domain-containing adapter protein 2-like 1 (BI2L1) and Heat shock protein 105 kDa (HSP105) increased with higher risk groups, consistent with reports linking both to aggressive tumor phenotypes[75–77]. Conversely, DNA damage-binding protein 2 (DDB2) and MAPK3 (MK03) decreased with increasing risk. Both proteins have been implicated in tumor-suppressive DNA damage and signaling pathways, although their roles in RCC remain unexplored[78–81].

## Conclusion and Clinical Implications

To our knowledge, this is the largest integrative and comparative MS imaging and proteomics study of RCC tumor subtypes ccRCC, chRCC, pRCC and OC for a representative number of patients within each group. High-throughput peptide profiling by MALDI-MSI with 541 TMA cores acquired in less than 48 hours differentiated RCC subtypes by their tryptic peptide mass profiles. A random forest classifier discriminated RCC subtypes with 100% accuracy at the patient level, representing first-in-class performance. Our optimized proteomics analysis of clinical RCC tumor microarray specimen quantified over 5000 proteins, distinguished common RCC subtypes, and identified a panel of candidate diagnostic and prognostic protein biomarkers.

We confirmed LAMP2 and UQCRC2 for discriminating the clinically challenging malignant chRCC from benign OC. ccRCC exhibit low levels of ATM, SETD3, CDK18 and high levels of CDK6 that were all associated with increased risk of relapse. Increased levels of BI2L1 and HS105 and reduced levels of MK03 and DDB2 correlated with an increased metastasis risk. These discoveries may aid urologists and pathologists in stratifying the RCC tumor spectrum to the benefit of patients.

Limitations of study: The speed and simplicity of peptide profiling by MALDI MS imaging comes at the cost of analytical depth as no peptides are sequenced and identified. Thus, the *m/z*-values of peptides in the MSI spectra are not annotated by the corresponding peptide or cognate protein identity. Peptide mass profiles suffice for accurate RCC tumor classification, as shown here, but does not allow for specific biomarker discovery by MSI based peptide profiling of cancer tissue. To complement the MALDI MSI experiments we also performed quantitative proteome analysis of each TMA core by LC-MS with the aim to identify proteins within the tumor tissues and to reveal differentially expressed putative biomarker proteins.

## Data Availability

All data produced in the present study are available upon reasonable request to the authors

## Acknowledgements

AAT was financed by a PhD fellowship from University of Southern Denmark (SDU), Department of Biochemistry and Molecular Biology. We thank Tina Ravnsborg, PhD and Andrea Maria Lorentzen for their help with protein sample preparation and mass spectrometry data acquisition.

## Author contributions

A.A.T, O.N.J, M.R. and N.M designed and outlined the project. M.R., N.M. and L.L. provided clinical samples and patient metadata. A.A.T. performed all experiments and data analysis. A.A.T. and O.N.J wrote the manuscript with feedback from M.R. N.M and L.L.

## References

1. Bray, F., et al., Global cancer statistics 2022: GLOBOCAN estimates of incidence and mortality worldwide for 36 cancers in 185 countries. CA: A Cancer Journal for Clinicians, 2024. 74(3): p. 229–263.

2. Moch, H., et al., The 2022 World Health Organization Classification of Tumours of the Urinary System and Male Genital Organs—Part A: Renal, Penile, and Testicular Tumours. European Urology, 2022. 82(5): p. 458–468.

3. Rini, B.I., S.C. Campbell, and B. Escudier, Renal cell carcinoma. Lancet, 2009. 373(9669): p. 1119–32.

4. Jayson, M. and H. Sanders, Increased incidence of serendipitously discovered renal cell carcinoma. Urology, 1998. 51(2): p. 203–205.

5. Tickoo, S.K. and M.B. Amin, Discriminant nuclear features of renal oncocytoma and chromophobe renal cell carcinoma. Analysis of their potential utility in the differential diagnosis. Am J Clin Pathol, 1998. 110(6): p. 782–7.

6. Tickoo, S.K., et al., Ultrastructural observations on mitochondria and microvesicles in renal oncocytoma, chromophobe renal cell carcinoma, and eosinophilic variant of conventional (clear cell) renal cell carcinoma. Am J Surg Pathol, 2000. 24(9): p. 1247–56.

7. Amin, M.B., et al., Renal oncocytoma: a reappraisal of morphologic features with clinicopathologic findings in 80 cases. Am J Surg Pathol, 1997. 21(1): p. 1–12.

8. Azawi, N.H., et al., Recurrence rates and survival in a Danish cohort with renal cell carcinoma. Dan Med J, 2016. 63(4).

9. Blum, K.A., et al., Sarcomatoid renal cell carcinoma: biology, natural history and management. Nature Reviews Urology, 2020. 17(12): p. 659–678.

10. Sanchez, A., A.S. Feldman, and A.A. Hakimi, Current Management of Small Renal Masses, Including Patient Selection, Renal Tumor Biopsy, Active Surveillance, and Thermal Ablation. J Clin Oncol, 2018. 36(36): p. 3591–3600.

11. Wendler, J.J., et al., Small renal carcinoma: the “when” and “how” of operation, active surveillance, and ablation. Pol J Radiol, 2018. 83: p. e561–e568.

12. Prieto, D.A., et al., Mass spectrometry in cancer biomarker research: a case for immunodepletion of abundant blood-derived proteins from clinical tissue specimens. Biomark Med, 2014. 8(2): p. 269–86.

13. Metatla, I., et al., Neat plasma proteomics: getting the best out of the worst. Clinical Proteomics, 2024. 21(1): p. 22.

14. Creighton, C.J., et al., Comprehensive molecular characterization of clear cell renal cell carcinoma. Nature, 2013. 499(7456): p. 43–49.

15. Lih, T.M., et al., Integrated glycoproteomic characterization of clear cell renal cell carcinoma. Cell Rep, 2023. 42(5): p. 112409.

16. Clark, D.J., et al., Integrated Proteogenomic Characterization of Clear Cell Renal Cell Carcinoma. Cell, 2019. 179(4): p. 964–983.e31.

17. Casadonte, R. and R.M. Caprioli, Proteomic analysis of formalin-fixed paraffin-embedded tissue by MALDI imaging mass spectrometry. Nature Protocols, 2011. 6(11): p. 1695–1709.

18. Judd, A.M., et al., A recommended and verified procedure for in situ tryptic digestion of formalin-fixed paraffin-embedded tissues for analysis by matrix-assisted laser desorption/ionization imaging mass spectrometry. J Mass Spectrom, 2019. 54(8): p. 716–727.

19. Angel, P.M., K. Norris-Caneda, and R.R. Drake, In Situ Imaging of Tryptic Peptides by MALDI Imaging Mass Spectrometry Using Fresh-Frozen or Formalin-Fixed, Paraffin-Embedded Tissue. Curr Protoc Protein Sci, 2018. 94(1): p. e65.

20. Johnson, W.E., C. Li, and A. Rabinovic, Adjusting batch effects in microarray expression data using empirical Bayes methods. Biostatistics, 2007. 8(1): p. 118–27.

21. Aumailley, M. and N. Smyth, The role of laminins in basement membrane function. J Anat, 1998. 193 (Pt 1)(Pt 1): p. 1–21.

22. Bachmann, M., et al., Cell Adhesion by Integrins. Physiol Rev, 2019. 99(4): p. 1655–1699.

23. Warrick, H.M. and J.A. Spudich, Myosin Structure and Function in Cell Motility. Annual Review of Cell and Developmental Biology, 1987. 3(Volume 3, 1987): p. 379–421.

24. Saito, Y., et al., Aldolase A promotes epithelial-mesenchymal transition to increase malignant potentials of cervical adenocarcinoma. Cancer Sci, 2020. 111(8): p. 3071–3081.

25. Zhang, K., L. Sun, and Y. Kang, Regulation of phosphoglycerate kinase 1 and its critical role in cancer. Cell Commun Signal, 2023. 21(1): p. 240.

26. Hamabe, A., et al., Role of pyruvate kinase M2 in transcriptional regulation leading to epithelial-mesenchymal transition. Proc Natl Acad Sci U S A, 2014. 111(43): p. 15526–31.

27. Abe, K., et al., Hypoxia-induced oxidative stress promotes therapy resistance via upregulation of heme oxygenase-1 in multiple myeloma. Cancer Medicine, 2023. 12(8): p. 9709–9722.

28. Barone, G., et al., The relationship of CDK18 expression in breast cancer to clinicopathological parameters and therapeutic response. Oncotarget, 2018. 9(50): p. 29508–29524.

29. Joshi, V., S.R. Lakhani, and A.E. McCart Reed, NDRG1 in Cancer: A Suppressor, Promoter, or Both? Cancers (Basel), 2022. 14(23).

30. Han, Y., D. Liu, and L. Li, PD-1/PD-L1 pathway: current researches in cancer. Am J Cancer Res, 2020. 10(3): p. 727–742.

31. Savoy, R.M. and P.M. Ghosh, The dual role of filamin A in cancer: can’t live with (too much of) it, can’t live without it. Endocr Relat Cancer, 2013. 20(6): p. R341–56.

32. Jiang, L., et al., Prognostic value of Musashi 2 (MSI2) in cancer patients: A systematic review and meta-analysis. Front Oncol, 2022. 12: p. 969632.

33. Diegmann, J., et al., Immune escape for renal cell carcinoma: CD70 mediates apoptosis in lymphocytes. Neoplasia, 2006. 8(11): p. 933–8.

34. Ždralević, M., et al., Double genetic disruption of lactate dehydrogenases A and B is required to ablate the “Warburg effect” restricting tumor growth to oxidative metabolism. J Biol Chem, 2018. 293(41): p. 15947–15961.

35. Luan, H., et al., EHD2 overexpression promotes tumorigenesis and metastasis in triple-negative breast cancer by regulating store-operated calcium entry. Elife, 2023. 12.

36. Wei, S., et al., EHD2 inhibits the invasive ability of lung adenocarcinoma and improves the prognosis of patients. Journal of Thoracic Disease, 2022. 14(7): p. 2652–2664.

37. Eskelinen, E.L., Roles of LAMP-1 and LAMP-2 in lysosome biogenesis and autophagy. Mol Aspects Med, 2006. 27(5-6): p. 495–502.

38. Li, L., et al., The HEAT repeat protein HPO-27 is a lysosome fission factor. Nature, 2024. 628(8008): p. 630–638.

39. Wei, Y., et al., Crystal structures of human lysosomal EPDR1 reveal homology with the superfamily of bacterial lipoprotein transporters. Commun Biol, 2019. 2: p. 52.

40. Fernandez-Vizarra, E. and M. Zeviani, Mitochondrial complex III Rieske Fe-S protein processing and assembly. Cell Cycle, 2018. 17(6): p. 681–687.

41. Zhang, Y., et al., MAPRE3 as an epigenetic target of EZH2 restricts ovarian cancer proliferation in vitro and in vivo. Exp Cell Res, 2024. 435(1): p. 113913.

42. Hartmann, C.M., et al., The precursor of mitochondrial aspartate aminotransferase is imported into mitochondria faster than the homologous cytosolic isoenzyme with the same presequence attached. Biochem Biophys Res Commun, 1991. 174(3): p. 1232–8.

43. Tian, E., et al., Galnt11 regulates kidney function by glycosylating the endocytosis receptor megalin to modulate ligand binding. Proc Natl Acad Sci U S A, 2019. 116(50): p. 25196–25202.

44. Prajapati, K., et al., The FLRT3-UNC5B checkpoint pathway inhibits T cell-based cancer immunotherapies. Sci Adv, 2024. 10(9): p. eadj4698.

45. Lam, V.K., et al., Morphology, Motility, and Cytoskeletal Architecture of Breast Cancer Cells Depend on Keratin 19 and Substrate. Cytometry A, 2020. 97(11): p. 1145–1155.

46. Yagnik, G., et al., Highly Multiplexed Immunohistochemical MALDI-MS Imaging of Biomarkers in Tissues. Journal of the American Society for Mass Spectrometry, 2021. 32(4): p. 977–988.

47. Dill, A.L., et al., Multivariate statistical differentiation of renal cell carcinomas based on lipidomic analysis by ambient ionization imaging mass spectrometry. Anal Bioanal Chem, 2010. 398(7-8): p. 2969–78.

48. Lu, H.C., et al., Imaging Mass Spectrometry Is an Accurate Tool in Differentiating Clear Cell Renal Cell Carcinoma and Chromophobe Renal Cell Carcinoma: A Proof-of-concept Study. J Histochem Cytochem, 2020. 68(6): p. 403–411.

49. Zhang, J., et al., Mass Spectrometry Imaging Enables Discrimination of Renal Oncocytoma from Renal Cell Cancer Subtypes and Normal Kidney Tissues. Cancer Res, 2020. 80(4): p. 689–698.

50. Möginger, U., N. Marcussen, and O.N. Jensen, Histo-molecular differentiation of renal cancer subtypes by mass spectrometry imaging and rapid proteome profiling of formalin-fixed paraffin-embedded tumor tissue sections. Oncotarget, 2020. 11(44): p. 3998–4015.

51. Prade, V.M., et al., The synergism of spatial metabolomics and morphometry improves machine learning-based renal tumour subtype classification. Clin Transl Med, 2022. 12(2): p. e666.

52. Shankar, V., et al., Distinguishing Renal Cell Carcinoma From Normal Kidney Tissue Using Mass Spectrometry Imaging Combined With Machine Learning. JCO Precis Oncol, 2023. 7: p. e2200668.

53. Qu, Y., et al., A proteogenomic analysis of clear cell renal cell carcinoma in a Chinese population. Nature Communications, 2022. 13(1): p. 2052.

54. Ricketts, C.J., et al., The Cancer Genome Atlas Comprehensive Molecular Characterization of Renal Cell Carcinoma. Cell Rep, 2018. 23(1): p. 313–326.e5.

55. Courtney, K.D., et al., Isotope Tracing of Human Clear Cell Renal Cell Carcinomas Demonstrates Suppressed Glucose Oxidation In Vivo. Cell Metab, 2018. 28(5): p. 793–800.e2.

56. Durán, R.V., et al., Glutaminolysis activates Rag-mTORC1 signaling. Mol Cell, 2012. 47(3): p. 349–58.

57. Xiao, Y. and D. Meierhofer, Glutathione Metabolism in Renal Cell Carcinoma Progression and Implications for Therapies. Int J Mol Sci, 2019. 20(15).

58. Miess, H., et al., The glutathione redox system is essential to prevent ferroptosis caused by impaired lipid metabolism in clear cell renal cell carcinoma. Oncogene, 2018. 37(40): p. 5435–5450.

59. Nguyen-Ngoc, K.V., et al., ECM microenvironment regulates collective migration and local dissemination in normal and malignant mammary epithelium. Proc Natl Acad Sci U S A, 2012. 109(39): p. E2595–604.

60. Zhang, Y.L., et al., Roles of Rap1 signaling in tumor cell migration and invasion. Cancer Biol Med, 2017. 14(1): p. 90–99.

61. Davis, Caleb F., et al., The Somatic Genomic Landscape of Chromophobe Renal Cell Carcinoma. Cancer Cell, 2014. 26(3): p. 319–330.

62. Zhang, H., et al., HIF-1 inhibits mitochondrial biogenesis and cellular respiration in VHL-deficient renal cell carcinoma by repression of C-MYC activity. Cancer Cell, 2007. 11(5): p. 407–20.

63. Simonnet, H., et al., Mitochondrial complex I is deficient in renal oncocytomas. Carcinogenesis, 2003. 24(9): p. 1461–1466.

64. Rohan, S., et al., Gene expression profiling separates chromophobe renal cell carcinoma from oncocytoma and identifies vesicular transport and cell junction proteins as differentially expressed genes. Clin Cancer Res, 2006. 12(23): p. 6937–45.

65. Drendel, V., et al., Proteomic distinction of renal oncocytomas and chromophobe renal cell carcinomas. Clin Proteomics, 2018. 15: p. 25.

66. Conner, J.R., M.S. Hirsch, and V.Y. Jo, HNF1β and S100A1 are useful biomarkers for distinguishing renal oncocytoma and chromophobe renal cell carcinoma in FNA and core needle biopsies. Cancer Cytopathol, 2015. 123(5): p. 298–305.

67. Ng, K.L., et al., Utility of cytokeratin 7, S100A1 and caveolin-1 as immunohistochemical biomarkers to differentiate chromophobe renal cell carcinoma from renal oncocytoma. Transl Androl Urol, 2019. 8(Suppl 2): p. S123–s137.

68. Li, G.X., et al., Comprehensive proteogenomic characterization of rare kidney tumors. Cell Reports Medicine, 2024. 5(5).

69. Ren, W., et al., Low Expression of ATM Indicates a Poor Prognosis in Clear Cell Renal Cell Carcinoma. Clinical Genitourinary Cancer, 2019. 17(3): p. e433–e439.

70. Giacosa, S., et al., Cooperative Blockade of CK2 and ATM Kinases Drives Apoptosis in VHL-Deficient Renal Carcinoma Cells through ROS Overproduction. Cancers (Basel), 2021. 13(3).

71. Abaev-Schneiderman, E., L. Admoni-Elisha, and D. Levy, SETD3 is a positive regulator of DNA-damage-induced apoptosis. Cell Death Dis, 2019. 10(2): p. 74.

72. Uhlen, M., et al., A pathology atlas of the human cancer transcriptome. Science, 2017. 357(6352): p. eaan2507.

73. Goel, S., J.S. Bergholz, and J.J. Zhao, Targeting CDK4 and CDK6 in cancer. Nature Reviews Cancer, 2022. 22(6): p. 356–372.

74. Butt, E. and D. Raman, New Frontiers for the Cytoskeletal Protein LASP1. Front Oncol, 2018. 8: p. 391.

75. Park, J.S., et al., Gene Expression Analysis of Aggressive Clinical T1 Stage Clear Cell Renal Cell Carcinoma for Identifying Potential Diagnostic and Prognostic Biomarkers. Cancers (Basel), 2020. 12(1).

76. Pipatpanyanugoon, N., et al., BAIAP2L1 enables cancer cell migration and facilitates phospho-Cofilin asymmetry localization in the border cells. Cancer Commun (Lond), 2022. 42(1): p. 75–79.

77. Kai, M., et al., Heat shock protein 105 is overexpressed in a variety of human tumors. Oncol Rep, 2003. 10(6): p. 1777–82.

78. MacNeil, A.J., et al., MAPK kinase 3 is a tumor suppressor with reduced copy number in breast cancer. Cancer Res, 2014. 74(1): p. 162–72.

79. Zhao, R., et al., DDB2 modulates TGF-β signal transduction in human ovarian cancer cells by downregulating NEDD4L. Nucleic Acids Res, 2015. 43(16): p. 7838–49.

80. Becuwe, P., et al., Manganese superoxide dismutase in breast cancer: from molecular mechanisms of gene regulation to biological and clinical significance. Free Radic Biol Med, 2014. 77: p. 139–51.

81. Kattan, Z., et al., Damaged DNA binding protein 2 plays a role in breast cancer cell growth. PLoS One, 2008. 3(4): p. e2002.

