## Supplementary Information for "Mass spectrometry and machine learning for classification and molecular phenotyping of renal cell carcinoma and benign tumors"

**Supplementary Table 1:** Included patients' age and gender.

| | Male | Female | Age 20-40 | Age 40-60 | Age 60+ |
|---|---|---|---|---|---|
| **ccRCC** | 54 | 39 | 0 | 31 | 52 |
| **pRCC** | 16 | 3 | 1 | 7 | 11 |
| **chRCC** | 5 | 4 | 1 | 3 | 5 |
| **OC** | 3 | 8 | 2 | 3 | 6 |
| **uRCC** | 2 | 2 | 0 | 0 | 4 |
| **ccpRCC** | 1 | 1 | 0 | 0 | 2 |
| **Sum** | **81** | **47** | **4** | **44** | **80** |
| **Total** | **128** | | **128** | | |

**Supplementary Table 2:** Sprayer settings for trypsin and CHCA application with the HTX M3+ sprayer.

| Type | Solvent | $N_2$ pressure (psi) | Height (mm) | Temperature (°C) | Track spacing (mm) | Flow Rate (mL/min) | Velocity (mm/min) | Passes | Moving Pattern | Drying time (s) |
|---|---|---|---|---|---|---|---|---|---|---|
| **Trypsin:** Syringe pump | 50 mM ABC, 10 ACN, 1% OG | 9.5 | 40 | 30 | 2 | 0.030 | 750 | 8 | CC | 0 |
| **CHCA (5 mg/mL):** Pos: peptides, glycans | 90% ACN, 9.9% $H_2O$, 0.1% TFA | 10 | 40 | 75 | 2 | 0.100 | 700 | 8 | CC | 0 |

**Supplementary Table 3:** Search settings for DIA-NN 1.8.1.

| Setting | Value |
|---|---|
| FASTA digest for Library-free search | Yes |
| Deep-learning based prediction | Yes |
| Missed cleavages | 2 |
| Cysteine carbamidomethylation as fixed modification | No |
| Oxidation of methionine | Yes |
| Peptide length | 7-30 amino acids |
| Precursor charge range | 1 - 4 |
| Precursor *m/z* range | 300 - 1800 |
| Fragment ion *m/z* range | 200 - 1800 |
| Precursor FDR | 1 % |
| MS1 accuracy | Optimized automatically (≈ 13 ppm) |
| Match-between-runs | Enabled |
| Protein inference | Genes |
| Quantification strategy | Robust LC |
| Cross-run normalization | RT-dependent |
| Speed and RAM usage | Optimal results |

### Supplementary Figure 1

A

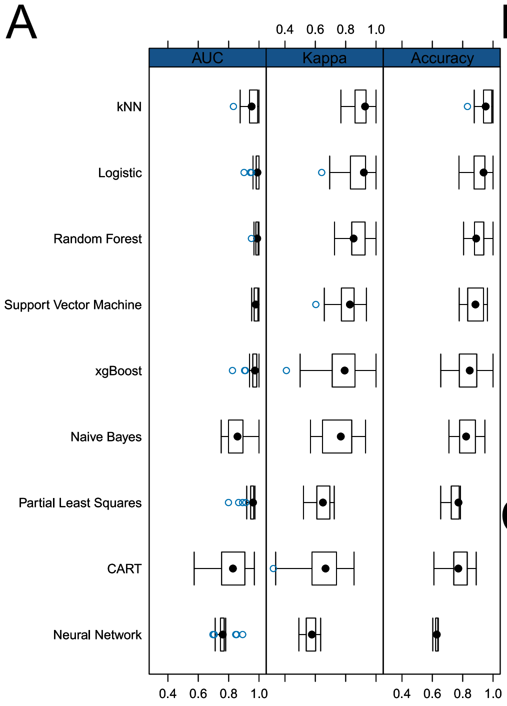


B

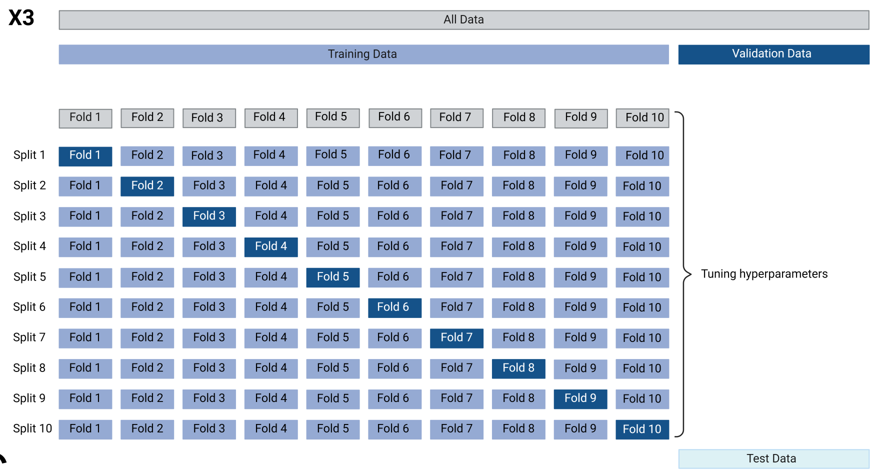


C

| Hyperparameter | Tuning range/values | |
|---|---|---|
| | Microproteomics | MALDI-MSI |
| Mtry | 12, 74, 1806 | 12, 17, 21, 26, 31 |
| Nodesize | 1-5 | 10,12,14,16,18,20,25,30,40,50 |
| Maxnodes | 5-15 | 5-15 |
| Ntree | 500, 600, 800, 1000, 1500, 2000 | 500,600,800,1000 |

**Supplementary Figure 1:** Supervised machine learning. (A) quality metrics for various machine learning models. x-axis represents values in the range of 0.4-1, y-axis depicts different machine learning models. Columns from left to right are area under the curve (AUC), kappa and balanced accuracy. (B) 10-fold 3x stratified repeated cross-validation was applied in the hypertuning process. Data was split 80:20 into training and test set. Training set was then used for tuning hyperparameters by using a 10-fold split applying the leave-one-out (LOOCV) principle. This process was repeated three times. (C) hyperparameters and their tuning range/values. Parameters mtry, nodesize, maxnodes and ntree was tested with different values or ranges depending on the dataset. The random forest model used pixels, i.e. individual mass spectra, as input and reached a performance of 95.6% mean balanced accuracy, a F1 score of 0.95 and a LogLoss of 0.22. The random forest model was trained on the RCC proteomics data with a similar split and class weights as for the MSI data. The classes ccRCC, chRCC, OC and pRCC were included in the model and the processed (filtered) was given as input.

### Supplementary Figure 2

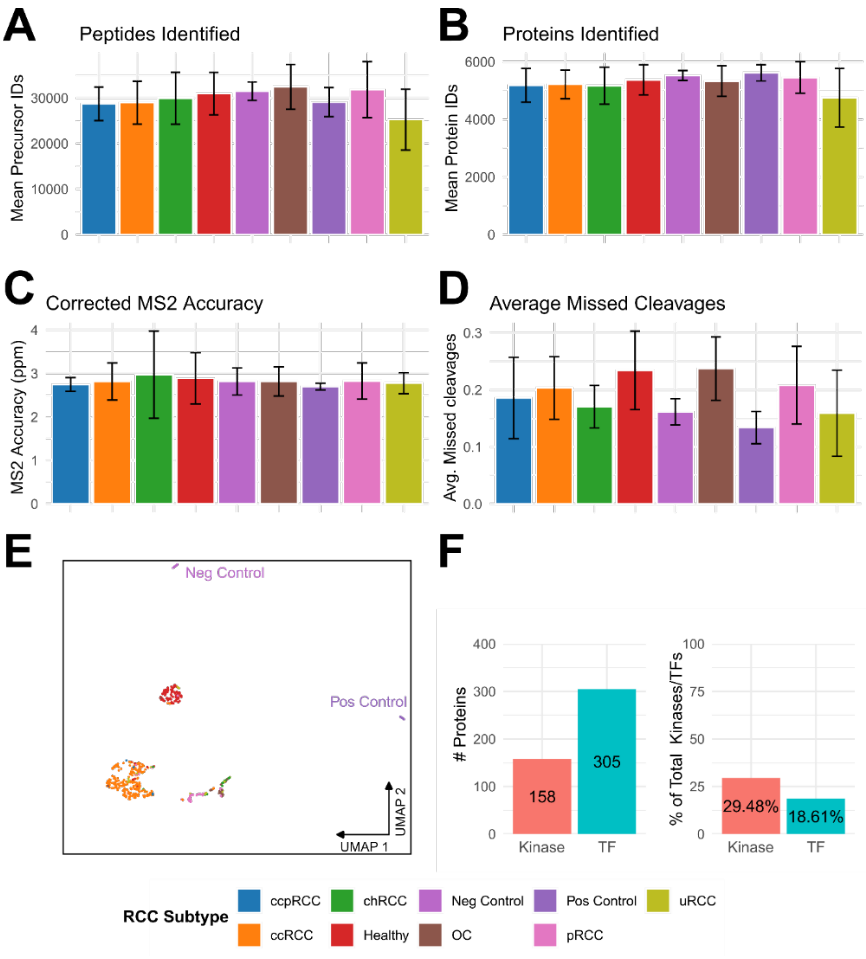


**Supplementary figure 2:** Quality control of microproteomics. (A) Barplot of precursors identified across RCC subtypes with error bars. y-axis represents mean precursor IDs. (B) barplot of proteins identified across RCC subtypes with error bars. y-axis represents mean protein IDs. (C) barplot of MS2 accuracy across RCC subtypes with error bars. y-axis represents MS2 accuracy in ppm. The mass accuracy of the peptide MS/MS spectra was 2.5-3 ppm which demonstrated an excellent performance of the LC-MS/MS setup. (D) barplot of average missed cleavages across RCC subtypes with error bars. y-axis represents average missed cleavages. An average missed tryptic cleavage site count of 0-1 reflects a high trypsin digestion efficiency and is a measure for the efficiency of our optimized *in situ* TMA sample preparation protocol. Too high numbers of missed cleavages would result decreased peptide coverage and impact quantification accuracy while introducing bias in the label-free quantification of peptides and proteins (E) UMAP of microproteomics data with each dot representing one sample. x-axis depicts first dimension of UMAP, y-axis represents the second dimension of UMAP. (F) barplots of count and percentage of total number of kinases and transcription factors (TFs). Barplot to the left depicts total number of kinases (lightred) and TFs (lightblue) identified. y-axis shows counts. Barplot to the right depicts the percentage out of all kinases (lightred) and TFs (lightblue) identified. y-axis shows percentage.

**Supplementary Table 4:** Molecularly defining KEGG terms associated with each subtype.

| | **Key KEGG Terms Defining RCC Neoplasms** |
|---|---|
| **ccRCC** | Glycolysis/Gluconeogenesis<br>Regulation of action cytoskeleton<br>HIF-1 signaling pathway |
| **pRCC** | No specific terms associated with the group |
| **chRCC** | Lysosome<br>Endocytosis<br>AMPK signaling pahway |
| **OC** | Oxidative Phosphorylation<br>Citrate cycle (TCA cycle)<br>Fatty acid metabolism |

### Supplementary Figure 3

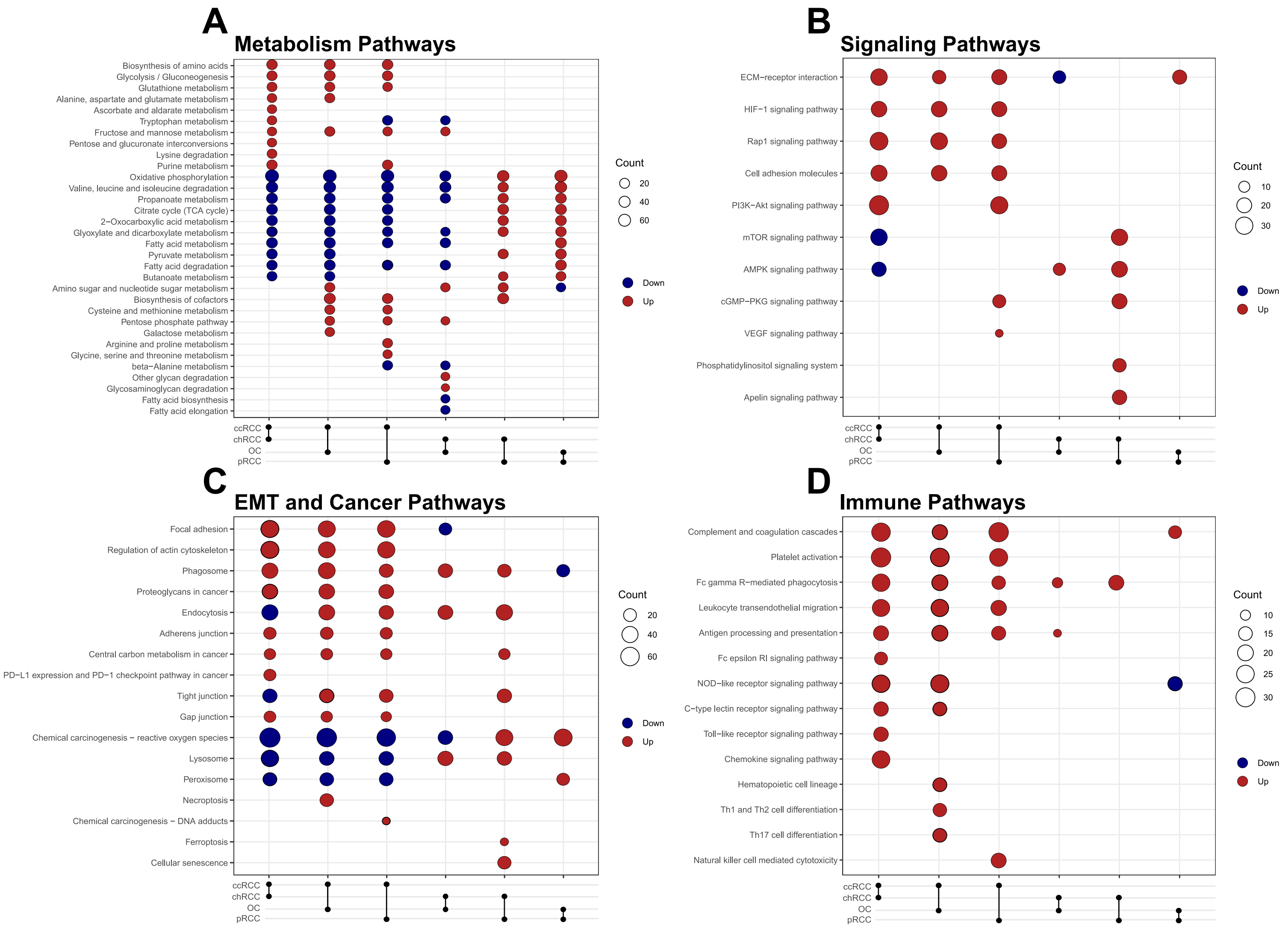


**Supplementary Figure 3:** Multiple testing of RCCs and OC refines molecular profiles generated by unsupervised analysis. (A) Dotplot of cluster comparison of enriched KEGG terms indexed to only include metabolism-related pathways. Count of proteins is depicted as circle size in legend, circle color indicates if KEGG term was up or down in the pairwise comparison with blue = down, red = up. x-axis shows the pairwise comparison with the top subtype indicating the first chosen for the comparison. y-axis depicts KEGG terms. (B) Dotplot of cluster comparison of enriched KEGG terms indexed to only include signaling-related pathways. Count of proteins is depicted as circle size in legend, circle color indicates if KEGG term was up or down in the pairwise comparison with blue = down, red = up. x-axis shows the pairwise comparison with the top subtype indicating the first chosen for the comparison. y-axis depicts KEGG terms. (C) Dotplot of cluster comparison of enriched KEGG terms indexed to only include EMT and cancer-related pathways. Count of proteins is depicted as circle size in legend, circle color indicates if KEGG term was up or down in the pairwise comparison with blue = down, red = up. x-axis shows the pairwise comparison with the top subtype indicating the first chosen for the comparison. y-axis depicts KEGG terms. (D) Dotplot of cluster comparison of enriched KEGG terms indexed to only include immune-related pathways. Count of proteins is depicted as circle size in legend, circle color indicates if KEGG term was up or down in the pairwise comparison with blue = down, red = up. x-axis shows the pairwise comparison with the top subtype indicating the first chosen for the comparison. y-axis depicts KEGG terms. ccRCC = clear cell renal cell carcinoma, chRCC = chromophobe renal cell carcinoma, OC = oncocytoma and pRCC = papillary renal cell carcinoma.

To confirm and strengthen our findings from the unsupervised KEGG enrichment, pairwise comparison and statistical testing followed by KEGG enrichment was performed. We initially split the dataset into separate dataframes for each RCC subtype (ccRCC, chRCC, pRCC, and OC). Welch's ANOVA was used to compare the protein expression across these groups, and subsequent post hoc Games-Howell tests were conducted to identify significant differences between RCC subtypes. Proteins exhibiting a >30% change in expression were selected for further analysis by KEGG pathway enrichment analysis for categories: metabolism pathways (S3A), signaling pathways (Figure S3B), EMT and cancer pathways (Figure S3C), and immune pathways (Figure S3D). ccRCC exhibited high levels of metabolic pathways associated with glucose metabolism and monosaccharides processing, alongside active biosynthesis of sulfur-containing amino acids, alanine, aspartate, and glutamate (Figure S3A). In contrast, mitochondrial processes such as the citrate cycle (TCA), oxidative phosphorylation (OXPHOS), fatty acid metabolism, and the degradation of valine, leucine, and isoleucine were downregulated in ccRCC. OC and chRCC displayed upregulation of mitochondrial-related pathways as compared to ccRCC and pRCC, with OC exhibiting stronger mitochondrial enrichment. Furthermore, chRCC exhibited increased glycan-degrading processes as compared to OC. For the signaling pathways (figure S3B) ccRCC showed upregulation of cytoskeletal remodeling and extracellular matrix reorganization, in addition to the HIF-1 signaling pathway. ChRCC displayed upregulation of the AMP-activated protein kinase (AMPK) pathway, distinguishing it from the other three RCC subtypes (figure S3B). When examining EMT and cancer pathways (figure S3C), ccRCC displayed high cell motility and migration potential, evidenced by upregulation of terms related to cytoskeletal regulation, focal adhesions, adherens junctions, and proteoglycans in cancer. It also exhibited activation of central carbon metabolism and phagosome-related processes, while reactive oxygen species (ROS) production was downregulated, suggesting a reliance on glycolysis in an active TME (figure S3C). ChRCC showed elevated expression of vesicle transport-related pathways, including peroxisome, lysosome, and endocytosis, indicating a prominent role of vesicular transport in chromophobe cancers. OC had higher ROS production than RCC subtypes, correlating with its distinct mitochondrial metabolic signature. ccRCC exhibited upregulation of multiple immune pathways including the complement system, antigen presentation and natural killer cells (figure S3D), whereas no significant trends were observed in immune terms for other subtypes.

In summary, our proteomics KEGG enrichment results from multiple testing agree with the findings from the unsupervised clustering KEGG enrichment. They support the hypothesis that ccRCC, chRCC, and OC exhibit distinct molecular profiles, as detailed by pathway enrichment analyses. These molecular differences suggest that while ccRCC adopts a glycolytic phenotype with a shift towards the Warburg effect and with high migration potential, chRCC and OC maintain distinct mitochondrial profiles which could indicates high energy demands and stress. In addition, chRCC has high levels of proteins associated with vesicle transport, likely reflecting a large vesicle load and upregulated transport processes.

**Supplementary Methods**

Sample Collection

Renal tumors from patients undergoing nephrectomy at Odense University Hospital, Odense between 2006 and 2018, were subtyped and evaluated by pathologists to select representative tumor areas with minimum 80% tumor cells (n = 3/patient) and adjacent healthy (n = 1/patient) tissue. Needle biopsies of renal masses were arranged in tissue microarrays (TMAs) for a total of 541 samples from 128 patients. The 541 needle biopsy cores had a diameter of 2 or 3 mm and were distributed over 26 tissue blocks with up to 30 cores/block. The renal cancer diagnoses included ccRCC (n = 251), pRCC (n = 52), chRCC (n = 27), OC (n = 31), clear cell papillary renal cell tumor (ccpRCC) (n = 4), unclassified RCC (uRCC) (n = 24), Healthy kidney (n = 116), negative control (placenta, n = 18, distinct tissue protein profile) and positive control (testis, n = 18, broad/diverse protein expression profile). More patient information can be found in supplementary table 1.

Sample Preparation and Protein Digestion

Each of the 26 formalin-fixed paraffin embedded (FFPE) TMA blocks were sectioned at 3 µm thickness using a microtome. Two adjacent thin sections were used for MSI analysis and LC-MS analysis, respectively. One such section was deposited onto an IntelliSlide (Bruker Daltonics) and prepared for MSI analysis. The adjacent section was deposited onto a microscopy glass slide and prepared for proteome analysis by LC-MS. Thus, two sets of 26 TMAs were prepared for analyses by MSI and LC-MS, respectively. The 2x26 TMA sections were initially incubated at 60°C for 1 h before serial washing with xylene (2x) for 5 min, 96% ethanol (EtOH) for 1 min, 70% EtOH for 30 sec, Carnoy's solution for 2 min, and tissue rehydration in an EtOH Milli-Q filtered water series: 96% 1 min, 70% 30 sec, Milli-Q 3 min x2[1-3]. Antigen retrieval was performed in Tris-HCL buffer adjusted to pH = 9 for 70 min at 95 °C[1, 2], followed by cool down to room temperature with a final wash in 10 mM ammonium bicarbonate for 5 min. All tissue slides were *in situ* trypsin digested with 20 µg of in-house methylated trypsin was dissolved in 970 µL to reach a concentration of 0.02 µg/µL (at an approximate on-tissue concentration of 10 ng/mm$^2$). The solvent used for trypsin was 50 mM ammonium bicarbonate, 10% acetonitrile and 1% octyl beta-d-glucopyranoside and deposited by a HTX M3+ sprayer followed by 2 hrs incubation at 37°C in a humidity chamber (a closed petri dish with wet Kimwipes[3]). HTX M3+ sprayer settings for trypsin deposition are listed in Supplementary Table 2.

For quantitative proteomics, peptides were extracted from each of the 541 cores by adding 2-3 µL of 50 mM ammonium bicarbonate onto the tissue core (corresponding to 1 µL per 1 mm tissue core diameter) and dispensing x10, three consecutive times, each time followed by transfer to a low-binding tube containing 20 µL of 50 mM ammonium bicarbonate. Samples were then acidified by addition of 2 µL of 10% trifluoroacetic acid and loaded onto Evotips according to the Evosep standard protocol (www.evosep.com) and stored at 5°C until analysis by LC-MS.

For MSI peptide profiling, the second set of 26 TMA slides of 541 cores were coated with α-cyano-4-hydroxycinnamic acid (CHCA) by a HTX M3+ sprayer. Details of MALDI matrix and spray settings are listed in Supplementary Table 2.

MALDI Immunohistochemistry (MALDI-IHC) Sample Preparation

MALDI-IHC was performed by using a primary antibody for protein binding and a secondary mass-tag conjugated antibody for detection by MALDI MS. Large tissue biopsies and TMAs from chRCC and OC patients were sectioned at 3 µm thickness on a microtome and deposited onto indium tin oxide (ITO) glass and IntelliSlides, respectively. All samples were prepared according to the AmberGen protocol for using conjugated antibodies. Slides were pre-melted by incubating at 60°C for 2 h prior to deparaffinization in xylene for 3x 5 min with a final wash of xylene:EtOH for 3 min. Following paraffin-removal a series of rehydration steps was done: 100% EtOH for 2x 2 min, 95% EtOH for 1x 3 min, 70% EtOH for 1x 3 min and 50% EtOH for 1x 3 min. A final wash was done in in tris buffered saline (TBS) buffer for 1x 10 min.

Antigen retrieval was performed in alkaline ethylenediaminetetraacetic acid (EDTA) buffer for 30 min at 95°C followed by cooldown for 30 min. Tissue blocking was performed to reduce background signal while minimizing non-specific binding. For tissue blocking the slides were initially washed in TBS for 10 min. Blocking buffer consisted of 2% (v/v) normal donkey serum, 5% (w/v) bovine serum albumin (BSA) in TBS-T (TBS supplemented with 0.05% (v/v) Tween-20). Tissue blocking buffer was added to immerse the samples, defined by a hydrophobic barrier pen. The tissue sections were blocked for 1 h at room temperature.

A primary working probe mixture (2% (v/v) normal donkey serum and 5% (w/v) BSA prepared in TBS supplemented with 0.05% (v/v) Tween20) was made by mixing blocking buffer with primary antibody to obtain a final antibody concentration of 2.5 µg/mL and passing through a filter unit for 1 min at room temperature and 21.000 g. The blocked tissue sections were place upright against a vertical surface angle to allow liquid to run off before applying the primary working probe mixture. Primary antibody incubation was done overnight at 4°C, followed by 3x 5 min washes with TBS-T. Immediately after, secondary antibody incubation was done with the secondary working probe mixture (blocking buffer with secondary anti-Rabbit conjugated antibody,0.5 µg/mL). Since the secondary mass-tagged antibodies are UV-sensitive, care had to be taken to avoid cleavage of the conjugated mass tag before antibody-antigen binding. Therefore steps ≥30 min had to be done in the dark. Secondary incubation was done similarly to blocking and primary antibody incubation but for 1 h at room temperature. After incubation the stained tissues were washed for 3x 5 min in TBS followed by 1x 10 sec. in 50 mM ammonium bicarbonate buffer. Final bulk washes for 3x 2 min in ammonium bicarbonate buffer was done before drying the samples in a desiccator for 90 min.

After the drying step the conjugated mass tags were cleaved by near UV-irradiation (~365 nm) for 10 min. Slides were then coated with CHCA matrix by a HTX M3+ sprayer. After matrix application each slide was inserted into a sealed humid chamber face down with a 5% isopranol solution as humidifying agent. This was done for 30 sec. to recrystallize the matrix and improve the signal-to-noise ratio (SNR) of the mass tags from the secondary antibody.

Proteome analysis by mass spectrometry

Quantitative proteome analysis was performed by nanoliter flow chromatographic separation of peptides and amino acids by LC-MS using an Evosep ONE LC (Evosep, Odense, DK) interfaced to a Bruker timsTOF Pro2 tandem mass spectrometer (Bruker Daltonics, Bremen, DE). After *in situ* trypsin digestion of sample cores within TMA sections, peptides were extracted [4-7] and analyzed using a preset preset 24 min LC gradient (60 samples per day) on the Evosep ONE LC. The mass spectrometer was operated in the diaPASEF mode, taking advantage of the acquisition method available on the timsTOF instruments, with optimized DIA windows optimized by py_diAID (ref) based on prior data-dependent acquisition experiments. The use of DIA-PASEF allows for nearly 100% duty cycle with ions being accumulated, separated and detected in packets simultaneously. The LC-MS injection order of the 541 core samples was randomized to reduce technical variation. Human protein sequence database searching was performed by DIA-NN v.1.8.1 (settings in Supplementary Table 3) using the library-free mode and the human proteome as input to generate an *in silico* library with decoys. Quantification was performed in DIA-NN using the MaxLFQ algorithm[8].

The quantified data was filtered with a false discovery rate (FDR) of 1% from the MaxLFQ algorithm and analyzed in R v. 4.3.3. Preprocessing of the data included filtering for ≥ 2 unique peptides, k-nearest neighbor imputation with k = 5 while removing proteins with more than 80% missing values if they were not unique to a single subtype and found in 50% of the given samples for the unique renal subtype.

Peptide profiling and IHC by MALDI MSI

MALDI-MSI spectra were acquired at 100 µm spatial resolution at a data acquisition speed of 40-50 cores per hour, on average. MALDI-MSI is well suited for the TMA format for rapid screening as the speed is limited only by the changing of sample plates by the MSI instrument operator, and not by the instrument data acquisition time of 40-50 TMA cores/hour. Therefore, fitting as many representative specimens as possible on a single glass slide yields high throughput. To reduce sample preparation time and reduce non-specific cleavage we used 2 hrs incubation time for the tryptic digestion, as the enzyme efficiency declines rapidly with overnight digestion[2, 9]. Ten slides were prepared simultaneously in one working day, but up to 20 slides is achievable to increase the throughput.

MALDI MSI peptide profiling and MALDI-IHC were performed on a Bruker timsTOF fleX/MALDI-2 tandem mass spectrometer in the positive ion mode. Data acquisition was optimized for *m/z* 600-3500 to detect tryptic peptides, with spatial resolution (pixel size) of 100 µm. This allowed for rapid analysis of all 541 samples in less than 48 hours of instrument runtime. The acquired imaging datasets (26 batches) were imported into SCiLS lab v. 2023a, merged into one master data file, converted to the .imzML format and imported into R at 10 ppm mass accuracy (manually confirmed in SCiLS lab by checking the mass resolution across the peaks for the combined dataset with 26 batches included). The Cardinal R package was used for data preprocessing, including total ion count (TIC) normalization, peak picking with a SNR of 3, peak alignment with 0.5 *m/z* tolerance and peak binning. Finally, the data was deisotoped with a 6 parts per million (ppm) tolerance and Pearson correlation coefficient threshold of 0.85. The final dataset contained a total of 866 features (i.e. peptide *m/z* values) in the range *m/z* 600-3500.

MALDI-IHC measurements were done on the same Bruker timsTOF fleX/MALDI-2 platform operated in the positive ion mode. Data acquisition was optimized for *m/z* 1000-2000 to detect the mass tag at *m/z* = 1139.61. A spatial resolution of 20 µm was used for the large biopsies while the TMA slides were acquired at a spatial resolution of 50 µm. The acquired imaging datasets were imported into SCiLS lab v. 2023a for ion image generation. Each TMA MSI experiment was converted into .imzML format and imported into R at 10 ppm mass accuracy and normalized to the TIC.

Statistical data analysis

Statistical analysis was performed in R v. 4.3.3. Unsupervised data analysis was used to identify features within the dataset, independent of diagnosis and tumor subtype. This included k-means

clustering of quantified proteins and dimensionality reduction by uniform manifold approximation projection (UMAP).
Comparative analysis of datasets from tumor subtypes was achieved by variance analysis (ANOVA) for RCCs and OCs while adjusting for multiple testing. For both the unsupervised analysis and the statistical testing, Kyoto Encyclopedia of Genes and Genomes (KEGG) enrichment analysis was performed to obtain cellular and biochemical insights of molecular networks in RCCs and OCs.

Supervised machine learning was utilized to build high-performance classifiers. The random forest algorithm was chosen for tumor classification due to its high performance, low risk of overfitting due to multiple decision trees, and its built-in function for feature importance ranking. Each model was hypertuned (the hyperparameters were mtry, ntrees, maxnodes and nodesize) and cross-validated by k-fold cross-validation with k = 10 (see Supplementary Figure 1 for more details on each model). Utilizing a random forest model, we obtained a high mean balanced accuracy for the proteomics data for the test set. Since the model was able to distinguish between the renal masses with high precision it enabled us to identify diagnostic protein biomarkers by examining the weighted importance for each protein for the random forest prediction.

Protein levels for known cancer-associated proteins were compared in terms of relapse probability applying Cox regression and Kaplan-Meier curves for the large cohort of ccRCC patients. The ccRCC samples were split into their respective Leibovich risk groups[10] and statistically tested by ANOVA while adjusting for multiple testing. Cox regression and statistical testing allow for the discovery of candidate prognostic biomarkers: relapse markers by Cox regression, and metastasis risk markers by statistical testing.

For MSI peptide data, unsupervised data analysis served to differentiate between subtypes, statistical testing identified discriminative peptide signals (features), and supervised classification grouped the RCC subtypes.

To address potential TMA slide-to-slide batch variation for unsupervised analysis the dataset was first subjected to comBAT correction (manuscript in preparation) by Log2-transformation of *m/z* intensity data and scaling by centering by mean subtraction from each observation followed by standard deviation division.

For the MALDI-IHC data the intensities for chRCC and OC tissue cores were compared and visualized by violin plots.

**Supplementary References**


1. Casadonte, R. and R.M. Caprioli, *Proteomic analysis of formalin-fixed paraffin-embedded tissue by MALDI imaging mass spectrometry.* Nature Protocols, 2011. **6**(11): p. 1695-1709.
2. Judd, A.M., et al., *A recommended and verified procedure for in situ tryptic digestion of formalin-fixed paraffin-embedded tissues for analysis by matrix-assisted laser desorption/ionization imaging mass spectrometry.* J Mass Spectrom, 2019. **54**(8): p. 716-727.
3. Angel, P.M., K. Norris-Caneda, and R.R. Drake, *In Situ Imaging of Tryptic Peptides by MALDI Imaging Mass Spectrometry Using Fresh-Frozen or Formalin-Fixed, Paraffin-Embedded Tissue.* Curr Protoc Protein Sci, 2018. **94**(1): p. e65.
4. Wisztorski, M., et al., *Droplet-Based Liquid Extraction for Spatially-Resolved Microproteomics Analysis of Tissue Sections.* Methods Mol Biol, 2017. **1618**: p. 49-63.
5. Raghunathan, R., M.K. Sethi, and J. Zaia, *On-slide tissue digestion for mass spectrometry based glycomic and proteomic profiling.* MethodsX, 2019. **6**: p. 2329-2347.
6. Ryan, D.J., et al., *Protein identification in imaging mass spectrometry through spatially targeted liquid micro-extractions.* Rapid Commun Mass Spectrom, 2018. **32**(5): p. 442-450.
7. Möginger, U., N. Marcussen, and O.N. Jensen, *Histo-molecular differentiation of renal cancer subtypes by mass spectrometry imaging and rapid proteome profiling of formalin-fixed paraffin-embedded tumor tissue sections.* Oncotarget, 2020. **11**(44): p. 3998-4015.
8. Cox, J., et al., *Accurate proteome-wide label-free quantification by delayed normalization and maximal peptide ratio extraction, termed MaxLFQ.* Mol Cell Proteomics, 2014. **13**(9): p. 2513-26.
9. Lin, Z., et al., *Evaluation and minimization of nonspecific tryptic cleavages in proteomic sample preparation.* Rapid Communications in Mass Spectrometry, 2020. **34**(10): p. e8733.
10. Leibovich, B.C., et al., *A scoring algorithm to predict survival for patients with metastatic clear cell renal cell carcinoma: a stratification tool for prospective clinical trials.* J Urol, 2005. **174**(5): p. 1759-63; discussion 1763.
